# Association of plasma and CSF cytochrome P450, soluble epoxide hydrolase and ethanolamides metabolism with Alzheimer’s disease

**DOI:** 10.1101/2021.03.09.21252423

**Authors:** Kamil Borkowski, Theresa L. Pedersen, Nicholas T. Seyfried, James J. Lah, Allan I. Levey, Chadwick M. Hales, Eric B. Dammer, Colette Blach, Gregory Louie, Rima Kaddurah-Daouk, John W. Newman, Alzheimer’s Disease Metabolomics Consortium

**Affiliations:** West Coast Metabolomics Center, Genome Center, University of California Davis, Davis, CA 95616, USA; Dept Food Science and Technology, University of California - Davis, Davis, CA 95616, USA; Department of Biochemistry, Emory University School of Medicine, Atlanta, GA 30322, USA; Department of Neurology, Emory University, Atlanta, GA, 30329 USA; Duke Molecular Physiology Institute, Duke University, Durham, NC, 27708 USA; Department of Psychiatry and Behavioral Sciences, Duke University, Durham NC 27708, USA; Department of Psychiatry and Behavioral Sciences, Duke Institute for Brain Sciences and Department of Medicine, Duke University, Durham NC 27708, USA; Western Human Nutrition Research Center, United States Department of Agriculture - Agriculture Research Service, Davis, CA 95616, USA; Department of Nutrition, University of California - Davis, Davis, CA 95616, USA

## Abstract

Alzheimer’s disease shares inflammatory origin with cardiometabolic disorders. Lipid mediators, including oxylipins, endocannabinoids, bile acids and steroids are potent regulators of inflammation, energy metabolism and cell proliferation with well-established involvement in cardiometabolic diseases. However, their role in Alzheimer’s disease is poorly understood. In the current study we provide comprehensive analysis of plasma and CSF lipid mediators in a case-control comparison of patients with Alzheimer’s disease, utilizing a targeted quantitative mass spectrometry approach. In both plasma and CSF, we observed Alzheimer’s disease patients to have elevated components of cytochrome P450/soluble epoxide hydrolase pathway and lower levels of fatty acids ethanolamides, when compared to the healthy controls. Multivariate analysis revealed that circulating metabolites of soluble epoxide hydrolase together with ethanolamides are strong and independent predictors for Alzheimer’s disease. Both metabolic pathways are potent regulators of inflammation with soluble epoxide hydrolase being reported to be upregulated in the brains of Alzheimer’s disease patients. This study provides further evidence for the involvement of inflammation in Alzheimer’s disease and argues for further research into the role of the cytochrome P450/soluble epoxide hydrolase pathway and fatty acid ethanolamides in this disorder. Further, these findings suggest that a combined pharmacological intervention targeting both metabolic pathways may have therapeutic benefits for Alzheimer’s disease.

## Introduction

Risk factors for Alzheimer’s disease (AD) include cardiometabolic disorders, low-grade systemic inflammation and altered lipid and energy metabolism [1-3]. Lipid mediators are lipid derived signaling molecules that regulate both acute and low-grade systemic inflammation and energy metabolism along with other processes [4]. Circulating lipid mediators have been implicated in the pathogenesis of cardiometabolic diseases [5-7], however, their involvement in AD is still not well described.

Four important families of such lipid mediators readily detected in the circulation are the oxygenated polyunsaturated fatty acids (i.e. oxylipins), the endogenous cannabinoid receptor activators and their structural equivalents (i.e. endocannabinoids, including monoacylglycerols and ethanolamides), bile acids and steroids. Oxylipins are mainly oxygenated products of polyunsaturated fatty acids (PUFAs) generated *via* 4 main pathways: cyclooxygenases (COX), generating mainly prostaglandins; lipoxygenases (LOX), generating mainly hydroxy-fatty acids, including leukotrienes and lipoxins; cytochrome P450 (CYP) and soluble epoxide hydrolase (sEH), generating epoxy-fatty acids and dihydroxy fatty acids respectively [4], and reactive oxygen-mediated formation of prostanoids and hydroxy-fatty acids (REF). Products from these pathways exhibit both pro- and anti-inflammatory actions (REF). Endocannabinoids are mainly fatty acids esters and amides, which are ligands for the cannabinoid receptors CB1 and CB2, the transient potential vanilloid receptor TRPV1 and G-protein-coupled receptor GPR55 [8], all highly expressed in the central nervous system [9]. Endocannabinoids regulate energy metabolism [8] and are generally consider anti-inflammatory [10]. Bile acids (BA) are generated by the liver as primary bile acids and secreted into the gut to aid in lipid digestion, and they are further metabolized by the gut microbiome form secondary bile acids [11]. After reabsorption from the gut into the blood stream, BAs regulate energy metabolism with different potency between primary and secondary species [12].

Changes in circulating lipid mediators in relation to AD were previously reported. Particularly, several oxylipins of the acute inflammation pathway were reportedly elevated in AD [13, 14] and pro-resolving (quenching activated inflammatory singal) lipid mediators have been suggested as potential treatment for AD [15]. Specific changes in bile acids metabolism, including a decrease in primary and an increase in secondary metabolites were also observed in AD subjects [16] and differences in bile acids clearance for cholesterol pathway were reported in AD [17]. Notably, bile acids and some steroids manifest neuroprotective functions through activation of steroid receptors [18]. However, a comprehensive analysis of the AD related changes in circulating lipid mediators is lacking and reports of lipid mediators in cerebrospinal fluid (CSF) are minimal. In the current work we utilize plasma and CSF samples of AD patients and cognitively normal controls from the Emory Goizueta Alzheimer’s Disease Research Center (ADRC). Using a case control approach, we provide comprehensive analysis of AD-associated changes in both circulating and CSF lipid mediators, including major classes of oxylipins, endocannabinoids, bile acids and some steroids, covering multiple aspects of inflammatory cascades and regulators of energy metabolism. Moreover, to minimalize the bias caused by postprandial fluctuation in plasma lipids, the fasting state of opportunistically collected samples was estimated using a novel predictive model that uses levels of circulating lipid mediators [19].

## Materials and methods

### Subjects

All participants from whom plasma and CSF samples were collected provided informed consent under protocols approved by the Institutional Review Board at Emory University. Cohorts included the Emory Healthy Brain Study (IRB00080300), Cognitive Neurology Research (IRB00078273), and Memory @ Emory (IRB00079069). All protocols were reviewed and approved by the Emory University Institutional Review Board. All patients received standardized cognitive assessments (including Montreal Cognitive Assessment (MoCA)) in the Emory Cognitive Neurology clinic, the Emory Goizueta Alzheimer’s Disease Research Center (ADRC) and affiliated Emory Healthy Brain Study (EHBS) [20]. All diagnostic data were supplied by the ADRC and the Emory Cognitive Neurology Program. CSF was collected by lumbar puncture and banked according to 2014 ADC/NIA best practices guidelines. All CSF samples collected from research participants in the ADRC, Emory Healthy Brain Study, and Cognitive Neurology clinic were assayed using the INNO-BIA AlzBio3 Luminex assay at AKESOgen (Peachtree Corners, GA). AD cases and healthy individuals were defined using established biomarker cutoff criteria for AD for each assay platform [21, 22]. In total, plasma samples were available for 148 AD patients and 133 healthy controls and CSF samples were available for 150 AD patients and 139 healthy controls. Plasma and CSF sample collection overlap (both plasma and CSF collected at the same day) was 145 for AD group and 133 for the control group. Cohort information is provided in **Table S1**.

### Quantification of lipid mediators

Plasma concentrations of non-esterified PUFA, oxylipins, endocannabinoids, a group of non-steroidal anti-inflammatory drugs (NSAIDs) including ibuprofen, naproxen, acetaminophen, a suite of conjugated and unconjugated bile acids, and a series of glucocorticoids, progestins and testosterone were quantified in 50µL of plasma by liquid chromatography tandem mass spectrometry (LC-MS/MS) after protein precipitation in the presence of deuterated metabolite analogs (i.e. analytical surrogates) [23]. CSF analyses were performed with 100µL samples prepared as previously reported for the analyses of sweat [24] and analyzed as reported for plasma. All samples were processed with rigorous quality control measures including case/control randomization, and the analysis of batch blanks, pooled matrix replicates and NIST Standard Reference Material 1950 – Metabolites in Human Plasma (Sigma-Aldrich, St Louis, MO). Extraction batches were re-randomized for acquisition, with method blanks and reference materials and calibration solutions scattered regularly throughout the set. Instrument limits of detection (LODs) and limits of quantification (LOQs) were estimated according to the Environmental Protection Agency method (40 CFR, Appendix B to Part 136 revision 1.11, U.S. and EPA 821-R-16-006 Revision 2). These values were then transformed into sample nM concentrations by multiplying the calculated concentration by the final sample volume and dividing by the volume of sample extracted. A complete analyte list with plasma LODs and LOQs have been reported [23]. The majority of analytes were quantified against analytical standards with the exception of eicosapentaenoyl ethanolamide (EPEA), palmitoleoyl ethanolamide (POEA), and the measured PUFAs [i.e. linoleic acid (LA); alpha-linolenic acid (aLA); arachidonic acid (AA); eicosapentaenoic acid (EPA); docosahexaenoic acid (DHA)]. For these compounds, area counts were recorded, adjusted for deuterated-surrogate responses and the relative response factors were expressed as the relative abundance across all analyzed samples. Reported monoacylglycerols (MAGs) are the sum of 1- and 2-acyl isomers, due to isomerization during sample processing.

### Fasting state assessment and sample selection

Many of the CSF and plasma samples from AD patients were collected following additional research consent in the course of patients’ clinical evaluations. Lumbar puncture procedures were nearly all scheduled in the morning, but fasting was not mandated in these individuals. Therefore, fasting state of the samples was estimated using previously published predictive equation, based on the raw plasma levels of 12(13)-EpOME, GCDCA and NO-Gly [19]. Only subjects with the probability of the fasting state > 60% were used for the plasma analysis. All subjects were used to compare lipid mediators level in CSF. CSF was reported not to manifest postprandial lipid fluctuations [25], additionally, comparing predicted fated to predicted non-fasted AD subjects reveal minimal differences in only 2 metabolites (**Table S2**).

### Statistical analysis

All statistical tests were performed using JMP Pro 14 (JMP, SAS institute, Carry, NC). Prior to analysis, data were tested for outliers using the robust Huber M test and missing data were imputed using multivariate normal imputation for variables which were at least 75% complete. The imputed numbers constitute less than 3% of the data for both plasma and CSF. Imputation facilitated multivariate data analysis and non-imputed data were used for univariate approaches. Additionally, variables were normalized, centered, and scaled using Johnson’s transformation, with normality verification using the Shapiro-Wilk test. The difference between the control and the AD group were assessed using t-test with gender, age, and race as covariates. Additionally, two-way ANOVA was used to test for the gender x group and race x group interactions. In case of significant interaction, the group effect was tested separately for the interacting factor. Correlations between MoCA score and lipid mediators were assessed using Spearman’s rank order correlation, to account for non-linear associations. This analysis was performed using only AD subjects, stratified by the assessed fasting state for plasma. CSF samples were analyzed without fasting state stratification. Multiple comparison control was accomplished with the false discovery rate (FDR) correction method of Benjamini and Hochberg with a q =0.2 [26].

Predictive models for AD were prepared using a combination of bootstrap tree and stepwise logistic regression modeling. Prior to analysis, subjects were randomly split into training (70%) and validation (30%) cohorts. Variables most frequently appearing in the models were identified by bootstrap tree: Number of layers = 50; split per tree = 3, bootstrap sampling rate for variables and subjects = 1. A variable contribution scree plot was generated using variable rank and the likelihood ratio of chi-square. The scree plot was used to determine a likelihood ratio of chi-square cutoff value for variables contributing to the model. Selected variables were then subjected to stepwise logistic regressions. A stepwise analysis was performed with the maximal validation r^2^ as the model stopping criteria, or if an additional step increased the Bayesian information criteria (BIC). Variables selected by the sptepwise approach were then used to build the model using logistic regression. Metabolites that the model contribution p-value < 0.05 were excluded, to ensure the strongest model with the minimal number of predictors.

Partial least square discriminant analysis (PLA-DA) was used to integrate AD related differences in metabolite levels between plasma and CSF. PLS-DA model was built using the nonlinear iterative partial least squares algorithm with K-Fold variation method (k =7) and included 235 variables from plasma and CSF, including metabolite levels and informative metabolite ratios. For the clarity purpose, only variables with a variable importance in projection (VIP) score > 1.4 we displayed on the loading plot.

Correlation between CSF and plasma metabolites was assessed using Spearman’s rank order correlation.

## Results

### Fasting state assessment

Analysis of opportunistically collected samples brings a challenge of the unknown fasting state. The control cohort contained samples collected in the fasted state per ADRC and EHBS protocols, but the AD cohort included many who had no collected fasting state information and consist of samples collected in both fasted and non-fasted states. Therefore, to allow a direct comparison of the control and AD groups, we assessed the estimated subject fasting state using our previously published predictive model [19]. As expected, control group was predicted to contain mostly fasted subjects (**Figure S1**). Out of 133 control subjects, 105 (i.e. 79%) were predicted as fasted, while 17 (i.e. 12%) as non-fasted, with a probability of >60% and 11 (i.e. 8%) had a fasting state probability of <60%. Out of 148 AD subjects, 60 (i.e. 40%) were predicted as fasted and 81 (i.e. 55%) as non-fasted, with a probability of >60% and 7 (5%) had a fasting state probability of <60%. Fifty percent of detected metabolites manifested difference between predicted fasted and non-fasted AD subjects in plasma and only minimal differences were observed in CSF (**Table S2**).

### Cytochrome P450/soluble epoxide hydrolase metabolism is elevated in AD subjects

We compared plasma and CSF lipid mediator concentrations between the control and the AD groups, using only estimated fasted subjects with probability>60% for plasma. In plasma, we detected 42 oxylipins (85 measured), 5 PUFAs (5 measured), 17 endocannabinoids (22 measured), 3 NSAIDs (4 measured), 19 bile acids (23 measured) and 8 steroid hormones (8 measured). The mean values and p-values for t-tests and two-way ANOVA interactions for all detected metabolites are provided in **Table S3**. Plasma group-fold differences in the oxylipin, endocannabinoids and PUFAs, projected onto their metabolic pathway, are presented in **Figure 1**. The largest differences were observed in the long chain omega-3 PUFA metabolism. Both EPA and DHA enzyme derived mono-alcohols (5-LOX-derived 5-HEPE and 4-HDoHE and 12-LOX-derived 12-HEPE and 14-HDoHE) were lower (1.5-fold in average) in the AD group, when compared to the control. On the other hand, the sEH EPA metabolite 17,18-DiHETE, was 3-fold higher in the AD group. In the AD group, the AA pathway manifested lower levels of the COX derived prostaglandins PGF2α and PGD2 (1.6-fold average). Additionally, the AD group showed lower levels of acylethanolamides (1.5-fold in average) derived from dihomo-gamma-linolenic acid (DGLEA), AA (AEA), docosatetraenoic acid (DEA), DHA (DHEA) and oleic acid (OEA). Notable are also lower levels of autooxidation markers, particularly the EPA-derived 9-HEPE (2-fold), linoleic acid (LA) -derived TriHOMEs (1.65-fold) and AA–derived isoprostanes (1.3-fold) in AD group.

**Figure 1.**
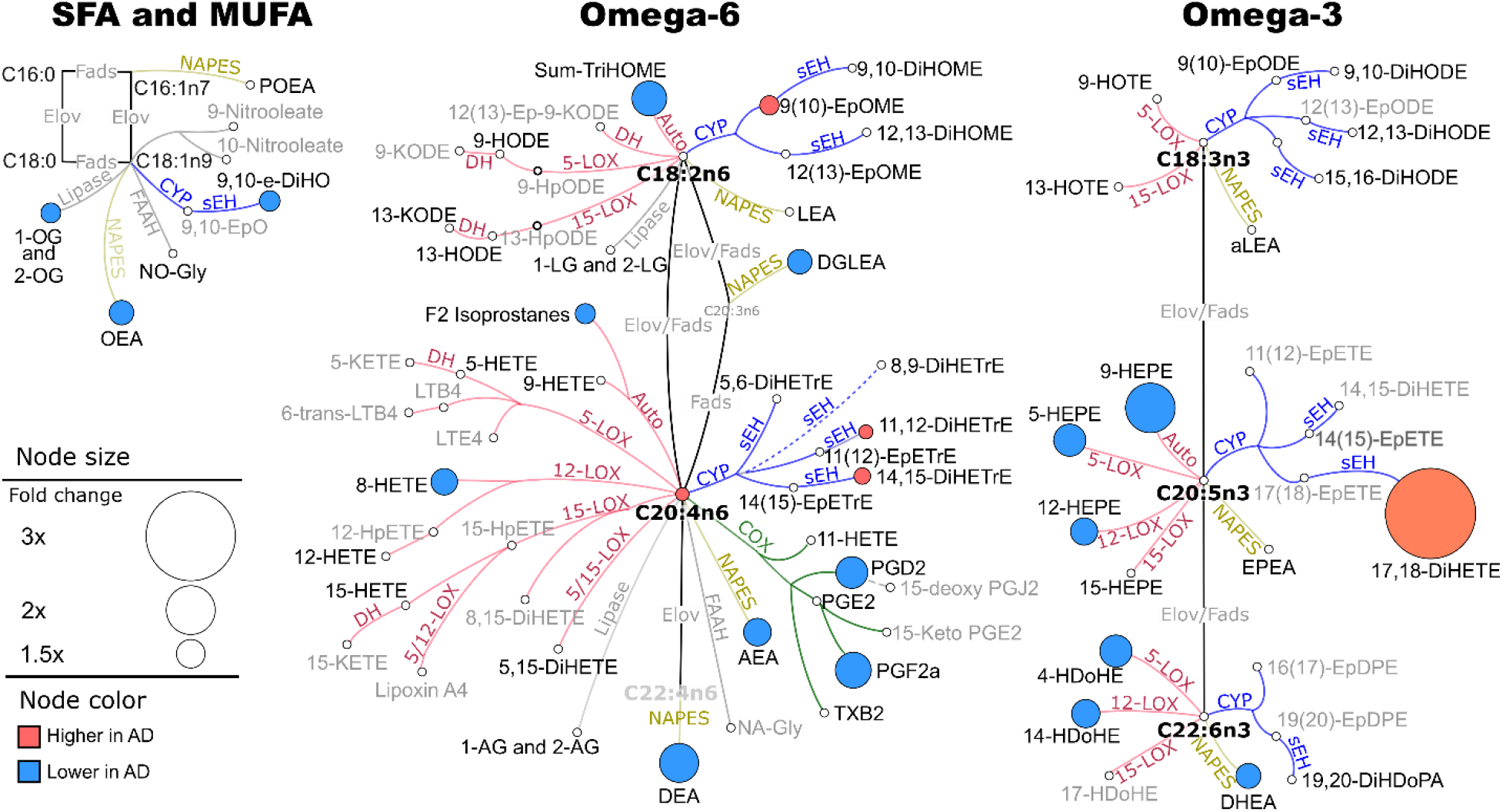
Plasma differences in fatty acids-derived oxylipins and endocannabinoids, between control and AD group, projected on their metabolic pathway. Only differences with the t-test p<0.05 and FDR correction at q=0.2 are shown. Network presents fatty acids metabolic pathway, including saturates and monounsaturates (SFA and MUFA) and omega 3 and omega 6 fatty acids with oxylipins and endocannabinoids synthesis pathway. Both detected (black font) and not-detected (gray font) metabolites are shown to visualize the coverage of the metabolic pathway in our targeted assay and facilitate data interpretation. Although, for the clarity of the figure, not all assay metabolites are displayed. The complete list of measured metabolites was previously reported [66]. Oxylipin metabolizing enzymes are colored by their class: red – lipoxygenase (LOX) and autoxidation pathway; blue – cytochrome p450 (CYP) epoxygenase; green – cyclooxygenase (COX); yellow – N-acylphosphatidylethanolamide-phospholipase D. Node size represents the-fold difference, and the color represents the directionality of the difference: orange – higher in AD; light blue – lower in AD. Key enzymes involved in metabolic step are abbreviated next to the edge. Fads – fatty acid desaturase; Elov – fatty acids elongase; sEH – soluble epoxide hydrolase; DH – dehydrogenase. Saturated and monounsaturated fatty acids were not measured in this assay and are indicated only to visualize precursors for measured oxylipins and endocannabinoids. Group means for all metabolites together with corresponding t-test p-values are provided in the **Table S3**.

Fewer lipid mediators were detected in CSF than in plasma. Detected CSF lipid mediators included 17 oxylipins, 5 PUFAs, 3 endocannabinoids, 14 bile acids and 6 steroids. The mean values and p-values for t-tests and two-way ANOVA interactions are provided in **Table S4**. CSF significant group-fold differences in the level of oxylipin, endocannabinoids and PUFAs, projected onto their metabolic pathway, are presented in **Figure 2**. In this matrix, the largest differences were observed in the LA CYP metabolic pathway, where both epoxy and dihydroxy FA, products of CYP and subsequent sEH metabolism, were higher in the AD group when compared to the control: epoxide average 1.5-fold; diol average 1.3-fold. All PUFAs from both omega-3 and omega-6 pathway were lower in the AD group, although the difference was only 1.2-fold on average. Additionally, AD group manifested 1.5-folds lower level of OEA and 1.3-fold lower level of the EPA-derived 14,15-DiHETE.

**Figure 2.**
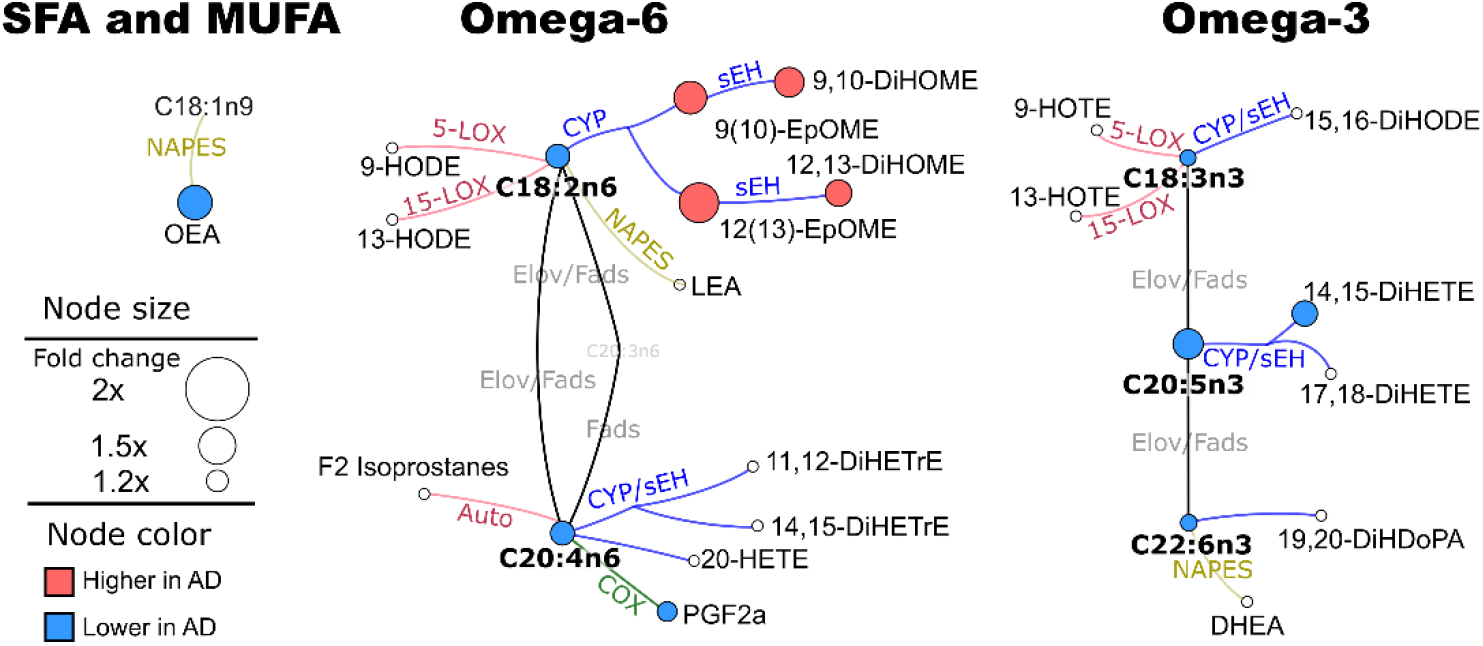
CSF differences in fatty acids-derived oxylipins and endocannabinoids, between control and AD group, projected on their metabolic pathway. Only differences with the t-test p<0.05 and FDR correction at q=0.2 are shown. Network presents fatty acids metabolic pathway, including saturates and monounsaturates (SFA and MUFA) and omega 3 and omega 6 fatty acids with oxylipins and endocannabinoids synthesis pathway. For the clarity purposes, only metabolites detected in CSF are shown. Oxylipin metabolizing enzymes are colored by their class: red – lipoxygenase (LOX) and autoxidation pathway; blue – cytochrome p450 (CYP) epoxygenase; green – cyclooxygenase (COX); yellow – N-acylphosphatidylethanolamide-phospholipase D. Node size represents the-fold difference, and the color represents the directionality of the difference: orange – higher in AD; light blue – lower in AD. Key enzymes involved in metabolic step are abbreviated next to the edge. Fads – fatty acid desaturase; Elov – fatty acids elongase; sEH – soluble epoxide hydrolase; DH – dehydrogenase. Saturated and monounsaturated fatty acids were not measured in this assay and are indicated only to visualize precursors for measured oxylipins and endocannabinoids. Group means for all metabolites together with corresponding t-test p-values are provided in the **Table S4**.

While few differences were observed in plasma and CSF bile acid levels between control and AD subjects, numerous differences were present in the specific bile acid ratios (**Table 1**). **Figure S2** shows bile acids metabolic pathway together with their median plasma levels to help understand the biological aspects of specific bile acid ratios. In plasma, the AD group was characterized by lower levels of cholic acid (CA), a product of the neutral bile acids synthesis pathway, while chenodeoxycholic acid (CDCA), a product of the acidic pathway was unchanged. This difference becomes even more pronounced when looking at the CA/CDCA ratio. On the other hand, the difference between neutral and acidic pathway was not present in downstream metabolites, when comparing the secondary unconjugated bile acids ratio, like deoxycholic acid/(lithocholic acid + ursodeoxycholic acid) (DCA/(LCA+UDCA)) or the most abundant primary conjugated derivatives glycocholic acid/glycochenodeoxycholic acid (GCA/GCDCA); **Table S3**. Of note, small differences between the neutral and acidic pathway were observed in the low abundance taurine conjugates of the secondary bile acids taurodeoxycholic/taurolithocholic acid (TDCA/TLCA). Difference in conjugation ratio (more conjugates than the substrate) was observed in the neutral synthesis pathway (GDCA/DCA and GCA/CA) but not in the acidic synthesis pathway. Differences between the neutral and acidic synthesis pathway were also observed in the conversion of the primary to secondary bile acids. The ratio of downstream products to their precursor in the neutral pathway was higher in the AD group in the case of DCA/CA, TDCA/CA and GDCA/CA, but not in parallel acidic pathway metabolites (i.e. LCA/CDCA, UDCA/CDCA).

**Table 1.** Differences in bile acids metabolites and their specific rations between control and AD group. Means are expressed in nM or as a ratio of the concentrations. Metabolites and their ratio are stratified by the metabolic affiliations. All tested bile acids and their ratios are presented in the **Table S3**.

| Metabolite | P value | Mean [95%CI] |  |
| --- | --- | --- | --- |
|  |  | Control | AD |
| Neutral vs acidic synthesis pathway |  |  |  |
| CA | 0.0321 | 27.4 [20.8 - 36.2] | 19.3 [13.6 - 27.4] |
| CDCA | 0.978 | 38.7 [29.2 - 51.5] | 46.7 [32.6 - 66.9] |
| CA/CDCA | 0.0008 | 0.707 [0.571 - 0.876] | 0.363 [0.271 - 0.487] |
| GCDCA/GDCA | 0.5 | 2.08 [1.66 – 2.61] | 1.97 [1.51 – 2.57] |
| TDCA/TLCA | 0.0114 | 1.33 [1.1 - 1.62] | 1.81 [1.49 - 2.21] |
| Conjugation, neutral synthesis pathway |  |  |  |
| GDCA/DCA | 0.0031 | 0.719 [0.604 - 0.856] | 1.05 [0.836 - 1.33] |
| TDCA/DCA | 0.1 | 0.0605 [0.0467 - 0.0783] | 0.0777 [0.0556 - 0.109] |
| GCA/CA | 0.0104 | 2.52 [1.86 - 3.39] | 4.13 [2.84 - 6.01] |
| TCA/CA | 0.076 | 0.506 [0.358 - 0.716] | 0.711 [0.434 - 1.16] |
| Conjugation, acidic synthesis pathway |  |  |  |
| GUDCA/UDCA | 0.7 | 0.546 [0.341 - 0.876] | 0.515 [0.334 - 0.793] |
| TUDCA/UDCA | 0.746 | 2.83 [1.93 - 4.17] | 2.04 [1.33 - 3.13] |
| GCDCA/CDCA | 0.491 | 8.15 [6.21 - 10.7] | 8.36 [5.68 - 12.3] |
| TCDCA/CDCA | 0.644 | 0.734 [0.53 - 1.01] | 0.687 [0.433 - 1.09] |
| TLCA/LCA | 0.961 | 0.282 [0.2 - 0.396] | 0.252 [0.182 - 0.347] |
| GLCA/LCA | 0.721 | 0.711 [0.53 - 0.955] | 0.749 [0.552 - 1.02] |
| Conversion of primary to secondary. Gut metagenome activity |  |  |  |
| DCA/CA | 0.0459 | 8.01 [5.8 - 11.1] | 12 [7.89 - 18.1] |
| TDCA/CA | 0.0056 | 0.469 [0.309 - 0.711] | 0.906 [0.517 - 1.59] |
| GDCA/CA | 0.0007 | 5.76 [4.01 - 8.27] | 12.5 [7.71 - 20.2] |
| LCA/CDCA | 0.451 | 0.66 [0.419 - 1.04] | 0.589 [0.42 - 0.825] |
| UDCA/CDCA | 0.691 | 1.39 [0.815 - 2.36] | 1.94 [1.23 - 3.07] |
| TUDCA | 0.0275 | 1.07 [0.78 - 1.48] | 1.92 [1.53 - 2.41] |

CSF manifested few differences in bile acids and their ratios. The AD group had 1.3-fold higher levels of GLCA and 1.4-fold higher level of T-a-MCA. Additionally, AD group had lower ratio of GCDCA/GLCA (1.3-folds, **Table S4**).

Of those measured, only a few steroid hormones showed different levels between AD and the control. In plasma, dehydroepiandrosterone sulfate (DHEAS) and progesterone were lower in AD group (1.9 and 1.7-folds respectively). Additionally, testosterone and the testosterone/progesterone ratio showed significant gender x group interaction. Females AD subjects showed 1.4-fold lower testosterone, when compared to females controls, but no differences were observed in males. On the other hand, the testosterone/progesterone ratio was 2-fold higher in AD male subjects compared male controls. Testosterone/progesterone ratio differences were not observed in females.

In CSF, only corticosterone showed a significant difference between AD and the control group, however, the magnitude of the-fold difference only ∼1.1.

### Relation between CSF and plasma AD markers

In the current study, matched plasma and CSF samples were collected, allowing an assessment of the relationships between metabolites in these pools. Spearman’s ρ rank order correlation between plasma and CSF lipid mediator levels are shown in **Table 2**. The associations were distinct by metabolite classes, with oxylipins showing only 2 of 15 significant correlations, while bile acids and steroids showing 14 of 18 significant correlations. Correlations within PUFA and PUFA ethanolamide were also apparent for the long chain omega 3 species (DHA EPA and DHA ethanolamide) but not others.

**Table 2.**
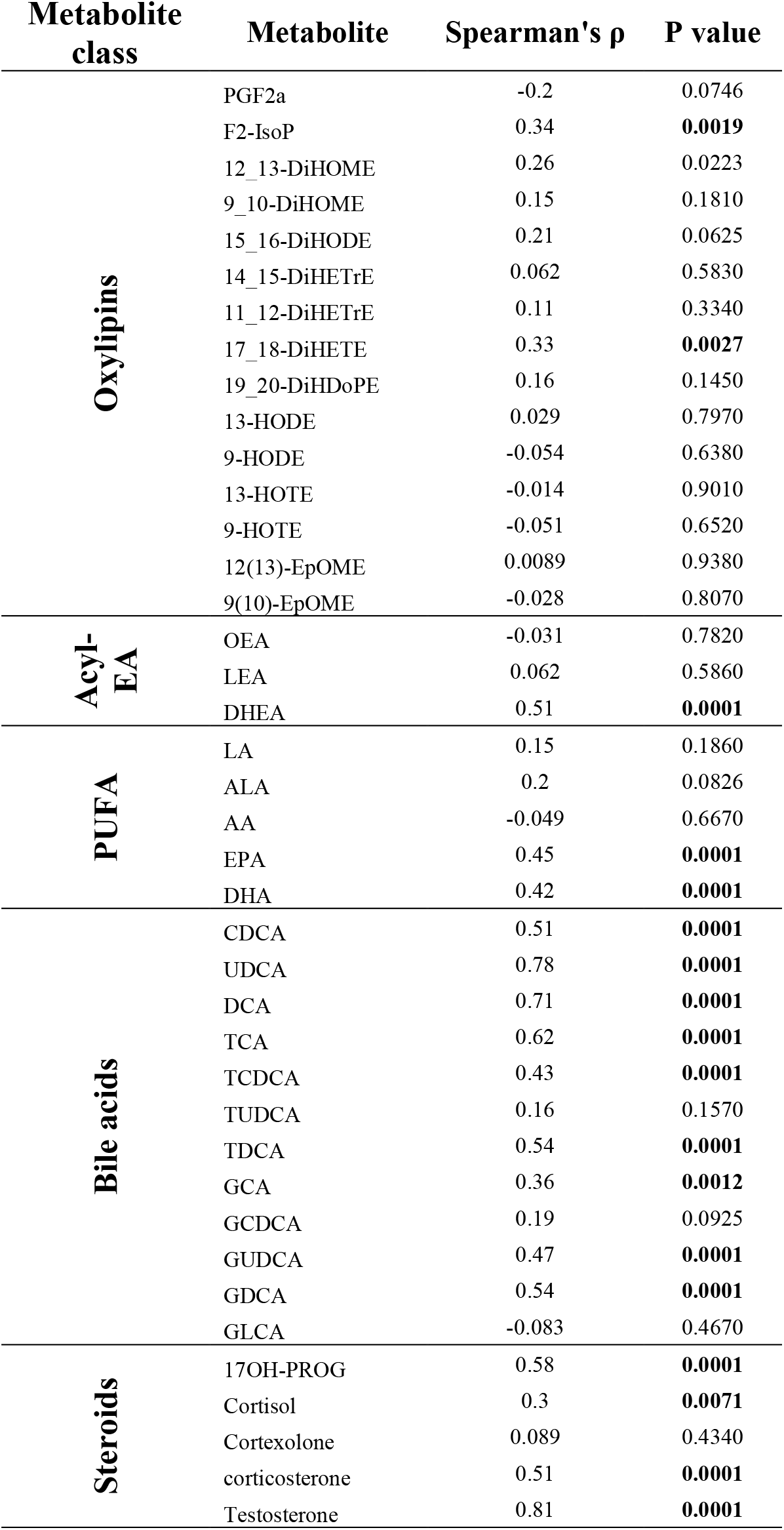
Spearmans rank order correlation between plasma and CSF metabolites. Significant p values are bolded.

Next, we used partial least square discriminant analysis (PLS-DA) to illustrate the relationship between plasma and CSF AD markers (**Figure 3**). The analysis showed that discrimination between control and AD was dominated by the plasma metabolites. Fifteen plasma metabolites (and their ratios) manifested variable importance in projection (VIP) score >1.4 compared to only 4 CSF metabolites. The discrimination between AD and the control group was characterized by higher plasma 17,18-DiHETE (VIP = 2.16) and CSF EpOMES (VIP = 1.95 and 1.58 for the 12(13) and 9(10) isoforms respectively) and lower levels of the acylethanolamide ratios including both DHEA/LEA and DEA/LEA in plasma and both plasma and CSF OEA/LEA. Plasma and CSF OEA/LEA manifest similar discriminatory power based on their proximity on the loading plot. On the other hand, plasma 17,18-DiHETE and CSF EpOMEs occupied distinct parts of the loading plot, suggesting distinct discriminatory properties. The VIPs for each metabolite are provided in the **Table S5**.

**Figure 3.**
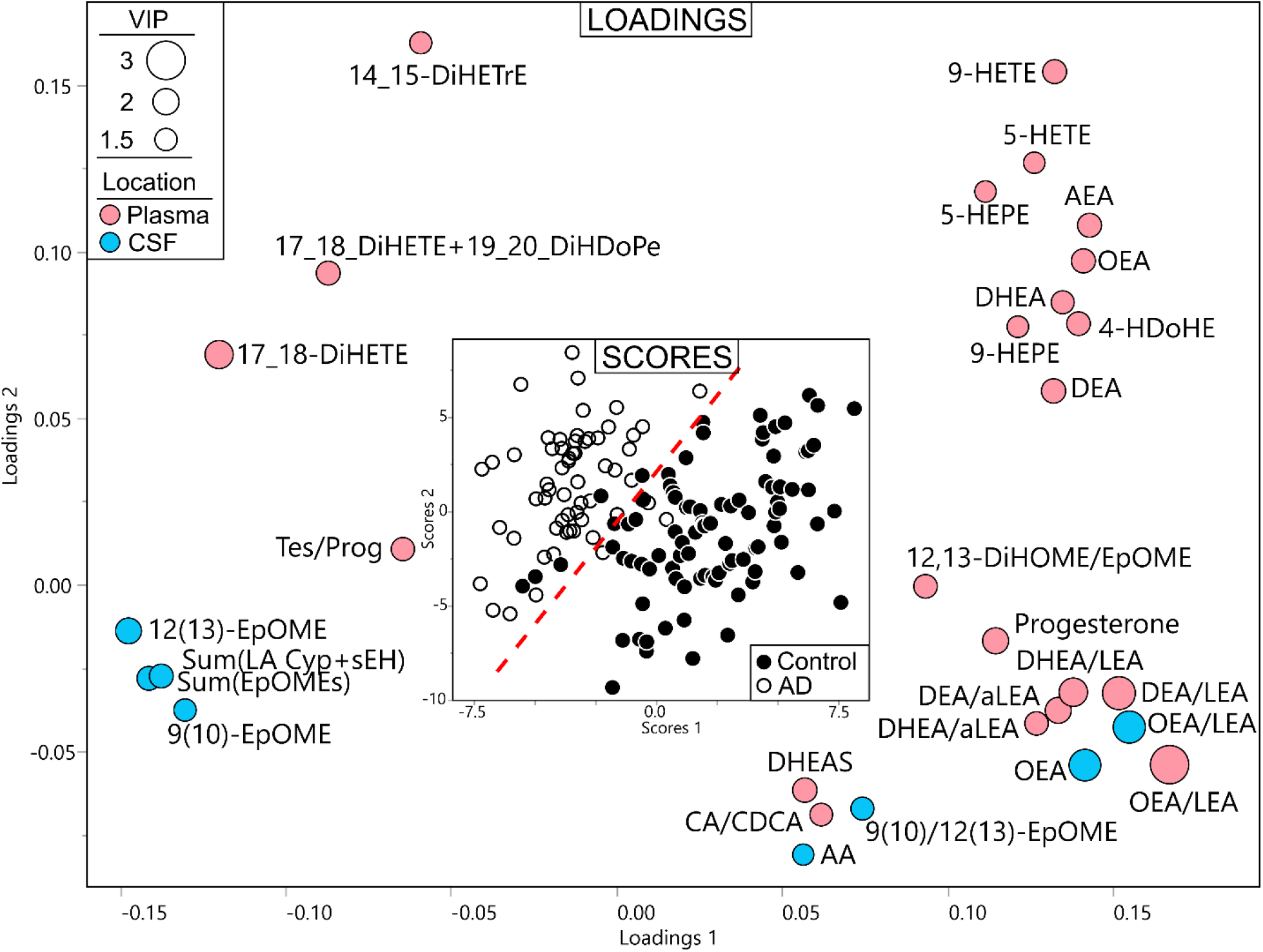
Relation between plasma and CSF predictors of AD. Partial least square discriminant analysis (PLS-DA) of AD vs control, utilizing metabolites from both plasma and CSF in predicted fasted samples. Treatment group discrimination is shown by the SCORES (inset) with a plain of discrimination indicated by dashed read line, while metabolites weighting in group discrimination are shown by the LOADINGS. Loading node color indicates metabolite origin (pink for plasma and blue for CSF). Loading node size indicates metabolite variable importance in projection (i.e. VIP). Analysis was performed with all measured metabolites, including specific ratios, but only those with VIP ≥1.4 are displayed for clarity purpose.

### Fatty acid ethanolamides and CYP/sEH metabolites are strong AD predictors in both plasma and CSF

We used predictive modeling to investigate how well plasma and CSF metabolites can report AD status. Plasma lipid mediators generated stronger models than those in CSF with area under the receiver operator characteristic curves (ROC AUC) of 0.924 vs. 0.824, with the two models consisting of distinct metabolites (**Figure 4)**. However, in both matrices, the strongest predictors belonged to the same two metabolic pathways, the acyl ethanolamides and CYP/sEH pathway. Plasma predictors included ethanolamides (OEA and DEA normalized to the LEA level), the 12,13-DiHOME/EpOME an indicator of sEH activity [27] and sEH metabolite of AA (14,15-DiHETrE). In CSF, the strongest predictors included OEA/LEA and the linoleate-derived epoxides 12(13)-EpOME and 9(10)-EpOME. When plasma and CSF markers were combined in predictive model efforts, the resulting model consisted uniquely of ethanolamides, including plasma long chain PUFA ethanolamides (DEA/LEA and DHEA/LEA) and CSF OEA/LEA. This model resulted in the ROC AUC of 0.889.

**Figure 4.**
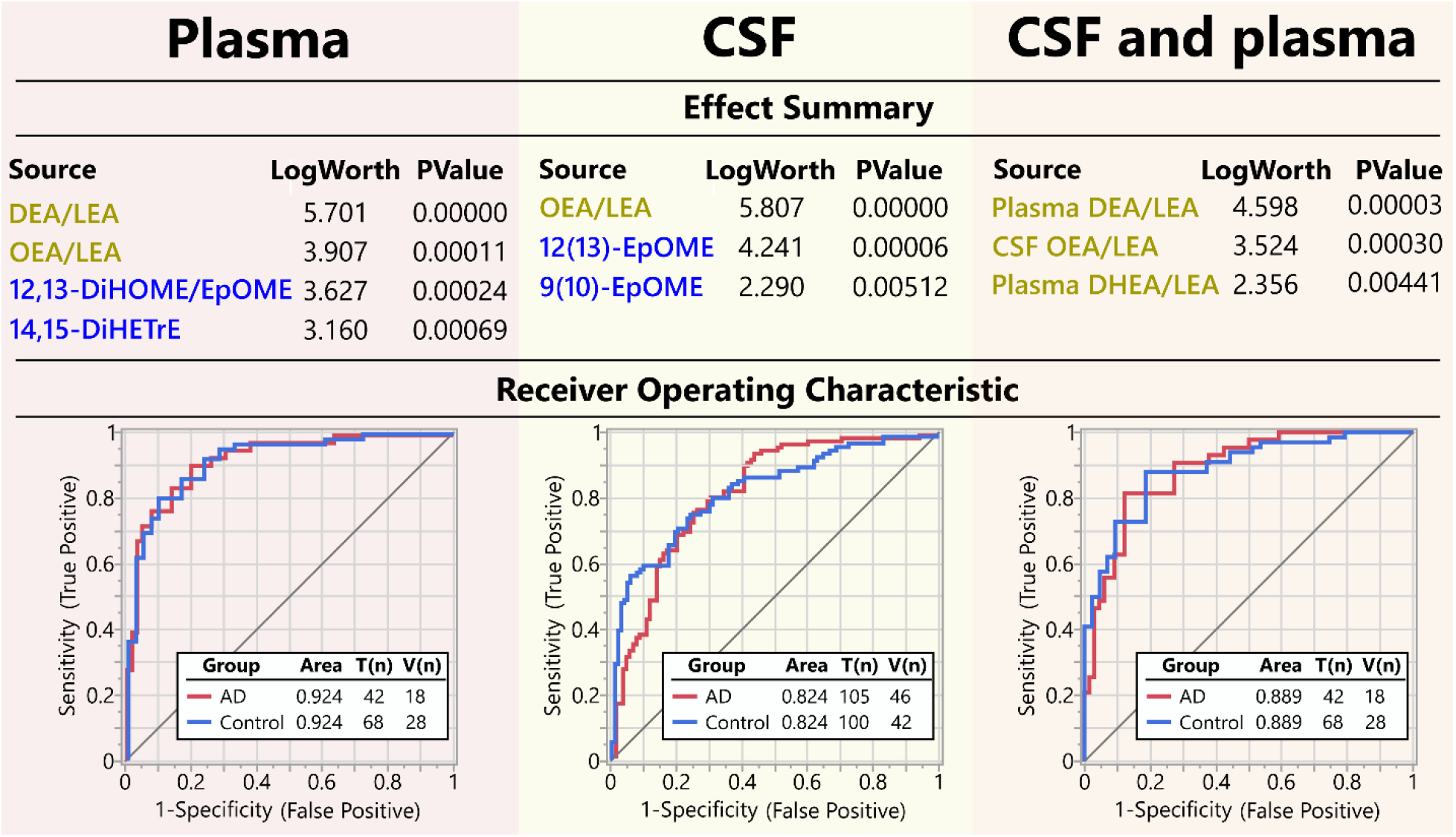
Predictive model for AD with plasma and CSF metabolites. Predictive model built independently for plasma (left) and CSF (middle) and plasma+CSF (right). Effect summery shows metabolite model components, sorted by their contribution to the model, with key pathway colored in yellow (fatty acids ethanolamides) and blue (cytochrome p450/soluble epoxide hydrolase pathway). Receiver operating characteristics (ROC) curve for the training set, together with the area under the curve (AUC) and the n for the training (T) and the validation (V) cohorts, are showed in the bottom panel. Metabolite selection through the stepwise logistic regression model together with validation cohort parameters are provided in the **Table S6**.

For all 3 models, ethanolamides OEA, DEA and DHEA were stronger predictors when used as a ratio to LA derived ethanolamide – LEA. LEA itself was not different between AD and the control group in either plasma or CSF (**Figure 1** and **Figure 2**) unlike OEA, DEA and DHEA. Therefore, LEA likely serves as a surrogate for the general acyl-ethanolamides level and adjustment of other ethanolamides by LEA lowers intra-individual variability.

### Lipid mediator–cognitive score associations in AD

The AD cohort is characterized by a high log(t-Tau/Αβ42) ratio and MoCA scores ranging from normal cognitive function to severe cognitive impairment (**Figure S3**). Taking advantage of the broad MoCA range, we investigated lipid mediator associations with cognitive function in this group pathological levels of t-Tau/Aβ42. Additionally, since the AD cohort was represented by subjects in both fasted and non-fasted states, we stratified the analysis by fasting state for plasma samples (**Table 3**). In the fasting state, PUFA oxidation markers, 5,15-DiHETE and 9-HETE were negatively associated with the MoCA score (although only 5,15-DiHETE passed FDR correction). 5,15-DiHETE can have enzymatic or autooxidative origin, whereas 9-HETE is a strictly an autooxidative product. 5,15-DiIHETE correlated with 9-HETE in fasted subjects with an R^2^=0.415 (n=60; p <0.001). In non-fasted AD subjects, a strong positive association between the MoCA score and EPA-derived ethanolamide (EPEA) as well as the levels of EPA and DHA were observed. Additionally, a positive correlation was detected between MoCA and the EPA-derived 17,18-DiHETE, the DHA-derived 14-HDoHE and the 18 carbon PUFAs (LA and ALA), however, these did not pass FDR correction.

In CSF, the linoleic acid derived epoxides 12(13)- and 9(10) -EpOMEs showed weak but significant positive correlations with MoCA (ρ >0.2, p <0.005; **Table 4**). Additionally, positive associations were observed between MoCA and DHA and DHA derived diol (19,20-DiHDoPE) and conjugated bile acids GCA (and the ratio of GCA to GDCA and GCDCA), TCDCA and the conjugated to unconjugated ratio for DCA and CDCA (GCA/GCDCA, GCDCA/CDCA and TCDCA/CDCA). However, only linoleic acid epoxides passed the FDR correction.

**Table 3.**
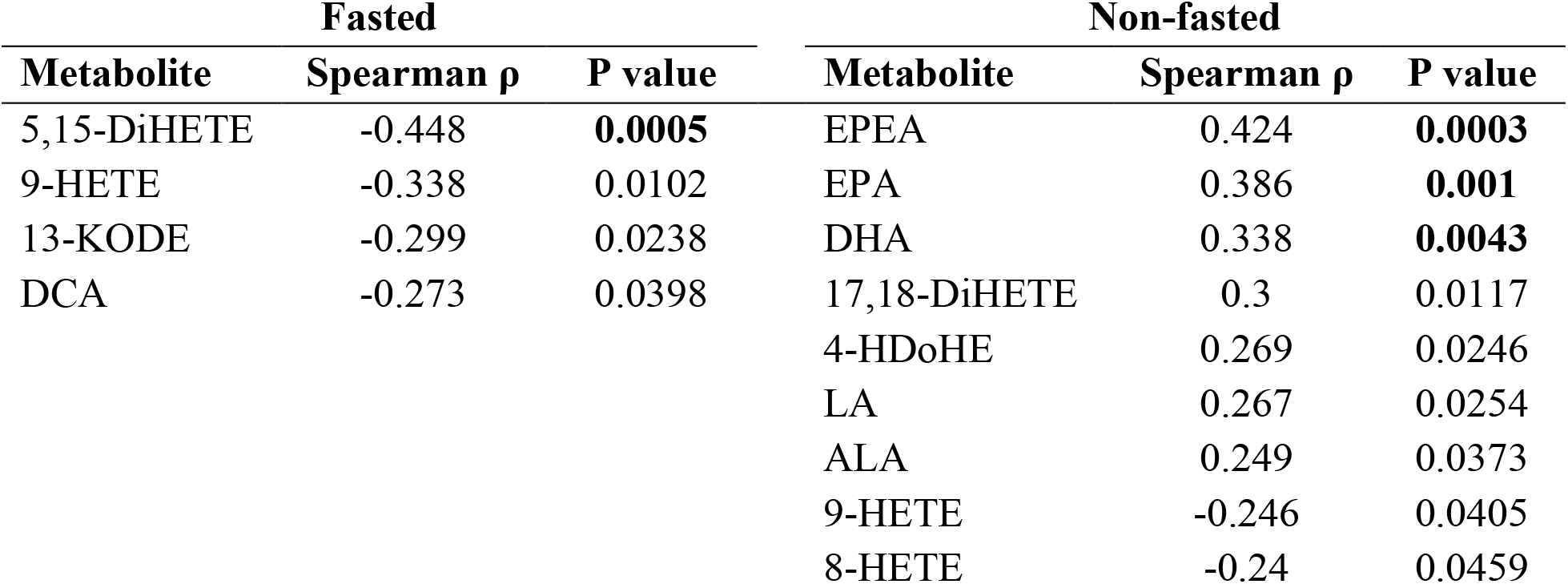
Spearman’s ρ rank order correlation between MoCA score and plasma lipid mediators of the AD patients. Analysis stratified by predicted fasted state. Only correlations with the p<0.05 are shown. P-values that passed FDR correction at q=0.2 are bolded.

**Table 4.** Spearman’s ρ rank order correlation between MoCA score and CSF lipid mediators of the AD patients. Only correlations with the p<0.05 are shown. P-values that passed FDR correction at q=0.2 are bolded.

| Metabolite | Spearman $\rho$ | P value |
| --- | --- | --- |
| 12(13)-EpOME | 0.279 | **0.0009** |
| 9(10)-EpOME | 0.24 | **0.0047** |
| 19,20-DiHDoPE | 0.21 | 0.0138 |
| GCA | 0.207 | 0.0152 |
| DHA | 0.203 | 0.0174 |
| GCA/GDCA | 0.202 | 0.0182 |
| GCDCA/CDCA | 0.19 | 0.026 |
| TDCA/DCA | 0.19 | 0.0261 |
| GCA/GCDCA | 0.18 | 0.0352 |
| TCDCA/CDCA | 0.172 | 0.0441 |
| TCDCA | 0.17 | 0.0468 |

## Discussion

Metabolic disruptions influencing vascular physiology, inflammation and energy metabolism have been reported to increase the risk of Alzheimer’s disease, however whether these changes are independent risk factors or how they may interact has not been well established. If novel biomarkers of AD can be identified within these domains, they could not only provide useful screening and risk assessment tools but may also provide insight into connections between metabolism and neurodegenerative diseases. To this end, we performed a comprehensive analysis of plasma and CSF lipid mediators, endogenous regulators of multiple processes including inflammation and energy metabolism and described their associations with AD and cognitive function. In the process, we identify clear differences between AD and healthy controls in two metabolic pathways, CYP/sEH and fatty acids-derived ethanolamides, and subtle differences in bile acids and steroids.

AD-associated differences in plasma bile acid were in agreement with previously reported analyses of The Religious Orders Study and the Rush Memory and Aging Project (ROS/MAP) cohort [16]. These included lower levels of CA and the CA:CDCA ratio in AD, suggesting that the neutral bile acid synthesis pathway could be affected in AD. There are few studies regarding the shift between neutral and acidic BA synthesis, one of them reporting an increase in neutral/acidic pathway products ratio in nonalcoholic steatohepatitis [28]. However, the biological relevance of this difference in terms of AD is yet to be determined. Interestingly, we previously reported associations between postprandial bile acids and cognition, with few associations in the fasting state [19]. Considering that the current manuscript focuses on AD related differences in the fasting state, further studies probing postprandial bile acid metabolism and AD are suggested. Interestingly, several bile acid and steroid differences in AD were gender specific. Few studies reports gender specific action of TUDCA and UDCA on ER stress markers in rodent model for the prion disease [29]. These findings further support the importance of gender focused approaches when investigating cholesterol-derived metabolism in the context of AD, as established with regards to the links between ApoE4 and AD risk in post-menopausal women [30].

With respect to fatty acid metabolism, our study identified substantial AD-associated elevations in CYP/sEH pathway products and lower levels of acylethanolamides in both plasma and CSF, although different elements of these pathways were affected in plasma and CSF. Oxylipin and endocannabinoid levels show no association between plasma and CSF, suggesting independent regulation of these pools. Likewise, CYP/sEH metabolites of plasma and CSF manifest distinct discriminatory power in PLS-DA models of AD. On the other hand, CSF and plasma acylethanolamides seem to manifest similar AD discriminatory power and can be substituted in the AD predictive model. These findings are consistent with previous reports implicating both CYP/sEH metabolites and acylethanolamides as important regulators of inflammation in neurodegenerative disorders [10, 31, 32].

In the CYP/sEH pathway, polyunsaturated fatty acids are converted to anti-inflammatory and vasodilating epoxy fatty acids by CYPs, which are further metabolized to pro-inflammatory and vasoconstricting diols by sEH, a process primarily recognized in cardiovascular disease [33]. In the current study, we found higher plasma level of the EPA sEH metabolite 17,18-DiHETE in AD patients, but only slight difference in the parallel AA metabolites were observed. Notably, EPA metabolites derived from LOX pathways were lower in AD patients, suggesting that the observed differences are not a result of differential omega-3 fatty acid intake, but rather specific enhancement in sEH-dependent EPA metabolism. Omega-3 sEH metabolites are particularly potent regulators of the cardiovascular system, especially blood vessel tone and vascular inflammation [34] and sEH inhibitors have been suggested to improve outcomes for both cardiovascular [35] and neurodegenerative diseases [36]. Clinical associations between cardiovascular disease and AD have been reported, where the regulation of a vascular tone and blood flow play a role in both pathologies [37]. Additionally, we previously reported plasma EPA sEH metabolites to be negatively associated with perceptual speed in cognitively normal subjects [19]. Therefore, the current findings further support involvement of vascular dysfunction in AD, perhaps through alterations to the blood brain barrier and vascular-related inflammatory signaling, with overlapping molecular mechanisms leading to cardiovascular and neurodegenerative pathologies. When considering these shifts in oxylipin profiles, it is important to remember that an epoxides reservoir is generated by esterification into phospholipid membranes [38], whereas diols are not readily reincorporated into the membranes and rapidly appear in the free pool and are actively excreted from cells [39]. Since tissue esterified lipid mediators were not evaluated in these samples, it is difficult to know whether the observe difference in EPA diol was due to an increased production of CYP/sEH metabolites or increased clearance of membrane bound EPA epoxides, and future studies are needed to resolve this issue.

In contrast to plasma, CSF showed higher levels of LA-derived epoxides, along with a moderate increase in LA-diols, but not CYP/EH metabolites of longer chain PUFAs. The source of the CSF metabolites is likely tied to the central nervous system, and linoleate-derived oxylipins have been identified as the dominant form in the developing rat brain [40]. Like long chain PUFAs, LA-derived epoxides and diols can also modulate vascular tone [41] and multiple studies point towards their cytotoxic and pro-inflammation nature. However most of these studies used concentrations greatly exceeding physiological levels [42] and cytotoxic effects were sEH-dependent, pointing towards LA diols as cytotoxic agents [43]. Interestingly, LA CYP/sEH metabolites elevated in the spinal tissue of burn victims were shown to activate the transient vanilloid receptor type 1 (TRPV1) [44]. Activation of the TRPV1 can rescue neuronal function from Aβ-induce impairment [45] and can alleviates cognitive and synaptic plasticity impairments in the *APP23/PS45* mouse model of AD [46]. Considering that acylethanolamides are also potent activators of the TRPV1 [47], increased LA CYP metabolites may compensate for the AD-related decrease in these ethanolamides. This hypothesis is supported by positive correlation of CSF LA-epoxides with the MoCA score in the AD patients, suggesting elevation of epoxy fatty acids in the central nervous system being potentially beneficial in AD. It is important to mention that the transportation of oxylipins in CSF is poorly understood. In plasma, the majority of oxylipins are transported as complex lipid esters in lipoproteins, with different lipoproteins manifesting distinct oxylipin compositions [48]. Therefore, the potential for lipoprotein-dependent oxylipin transport within CSF specific HDL particles [49] is particularly intriguing and warrants further investigation.

Together the CSF and plasma results implicate changes in both peripheral and central CYP/sEH metabolism in association with AD and cognitive impairment. These conclusions are consistent with previous reports of single nucleotide polymorphisms (SNPs) in the CYP2J2 promoter region that reduces gene expression by ∼50% that appears to increase the ApoE4-independent AD risk [50]. Several functional SNPs are also known to influence sEH activity and/or expression and influence disease risk [51]. Relevant to AD-associated pathologies, loss of function sEH mutations protect neurons from ischemia-induced death [52] and may alter the risk of vascular cognitive impairment [53]. Additionally, postmortem brains from human subjects with AD show higher sEH levels, when compared to the healthy controls, and sEH inhibitors can reverse microglia and astrocyte reactivity and immune pathway dysregulation in mouse AD models [31]. Additionally, brain sEH was positively associated with AD in a replicated protein wide-association study of AD [54]. Therefore, reducing sEH function appears to be protective, and supports sEH as a valuable therapeutic target for the treatment and investigation of neuro-inflammatory pathologies including AD.

Both plasma and CSF acylethanolamides were lower in AD, with both PLS-DA and predictive model identifying OEA as the strongest predictor of AD in the current cohort. Acylethanolamides are generally considered anti-inflammatory [55] and neuroprotective [56] and were previously implicated in neuroinflammatory processes [57]. Their neuroprotective action is mediated by activation of the CB1 and CB2 receptors [58] and TRPV1, involved in the acute and inflammatory pain signals in periphery [59]. Some acylethanolamides, like OEA, are also peroxisome proliferator-activated receptor (PPAR) α agonist [58] and regulate satiety and sleep with both central and peripheral anorexigenic effects. Notably, sleep disturbances themselves have been reported to be a risk factor for AD [60], and the identified reductions in CSF OEA would be consistent with such a physiological manifestation [61]. A recent study also suggested that the EPA-derived ethanolamide (EPEA) is a potential PPAR γ agonist [62], a transcription factor known for its neuroprotective and anti-inflammatory action [63]. Interestingly, non-fasting levels of EPEA showed positive association with MoCA in AD patients. This is in agreement with our previous findings of acylethanolamides in non-fasted individuals, including EPEA, being positively associated with perceptual speed in cognitively normal elderly individuals [19]. Literature provides conflicting results regarding both the levels of acylethanolamides in biological fluids, as well as the expression of CB1 and CB2 receptors in the context of AD [64]. Nevertheless, the body of literature suggests that exogenous cannabinoids are potent activators of the CB1 and CB2 receptors with potential therapeutic benefit for AD treatment, due to their neuroprotective and anti-inflammatory activity [65]. Our data suggest acylethanolamide biology is altered in relation to both AD pathology as well as cognition. However, future studies are needed to fully elaborate the role of these endocannabinoids in AD pathology.

In conclusion, the current study shows AD related differences in CYP/sEH and acylethanolamide metabolism observed in both plasma and CSF. Strong predictive and discriminant models suggest their potential as biomarkers of AD-associated metabolic disruptions. This further supports the contention that a combination therapy reducing sEH activity while increasing acylethanolamide tone by either promoting production or reducing degradation could be a more effective strategy than targeting either pathway independently in treating multifactorial inflammatory diseases like AD [31]. Important questions remain regarding the metabolic changes in the lipid mediators preceding pathological changes in tau and cognitive decline. We have previously reported that plasma sEH metabolites of the long chain omega-3 PUFA were negatively associated and PUFA ethanolamides positively associated with perceptual speed [19], mimicking the currently described AD related associations. While these data suggest early alterations in these important regulatory pathways, a comprehensive analysis of longitudinal metabolome changes in relation to cognition and tauopathies is warranted. Combining assessments of dietary, lifestyle and genetic factors promoting these metabolic changes offers the opportunity for novel risk factor discovery and the development of targeted preventive measures.

## Limitations

This study was conducted using opportunistically collected samples with the fasting state estimated using a previously developed predictive model [19] with an ∼17% inherent misclassification rate.

## Data Availability

Aggregated metabolomic data are provided as supplemental tables.

## Acknowledgements

Metabolomics data is provided by the Alzheimer’s Disease Metabolomics Consortium (ADMC) and funded wholly or in part by the following National Institute on Aging grants and supplements, components of the Accelerating Medicines Partnership for AD (AMP-AD) and/or Molecular Mechanisms of the Vascular Etiology of AD (M2OVE-AD): NIA R01AG046171, RF1AG051550, RF1AG057452, R01AG059093, RF1AG058942, U01AG061359, U19AG063744 and FNIH: #DAOU16AMPA awarded to Dr. Kaddurah-Daouk at Duke University in partnership with a large number of academic institutions. As such, the investigators within the ADMC, not listed specifically in this publication’s author’s list, provided data but did not participate in analysis or writing of this manuscript. A complete listing of ADMC investigators can be found at: https://sites.duke.edu/adnimetab/team/. Metabolomics data and pre-processed data are accessible through the AMP-AD Knowledge Portal (https://ampadportal.org). The AMP-AD Knowledge Portal is the distribution site for data, analysis results, analytical methodology and research tools generated by the AMP-AD Target Discovery and Preclinical Validation Consortium and multiple Consortia and research programs supported by the National Institute on Aging. Additional support was provided by Emory ADRC P30 AG066511 awarded to Allan I. Levey, Emory EHBS R01 AG070937 awarded to James J. Lah and USDA Intramural Projects 2032-51530-022-00D and 2032-51530-025-00D awarded to John W. Newman. The USDA is an equal opportunity employer and provider.

## Supplemental figures

**Figure S1.**
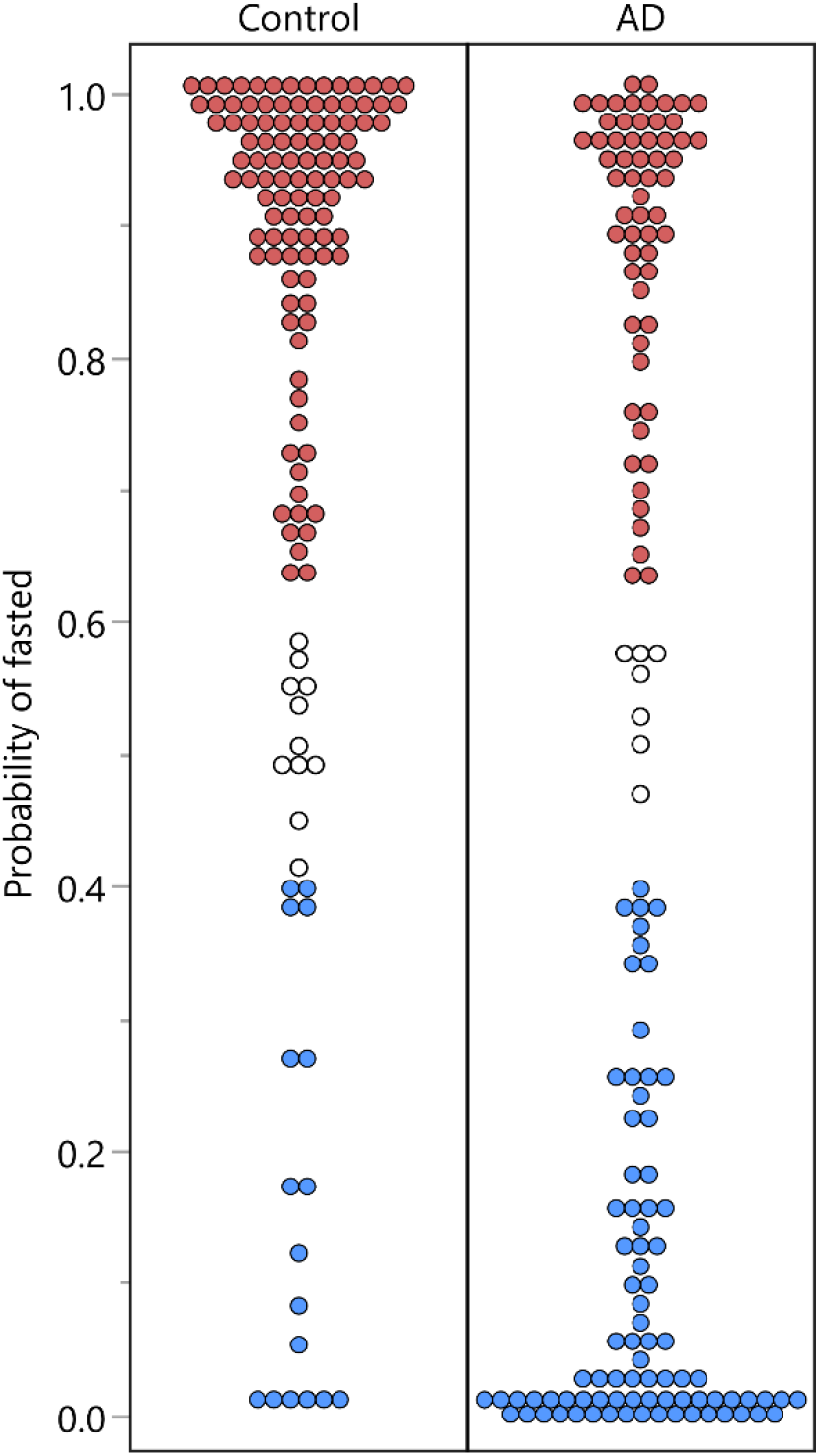
Distribution of subject’s probability of being in a fasted state in the control and AD groups. Colors shows 60% probability cutoffs for: red – fasted; blue – non-fasted.

**Figure S2.**
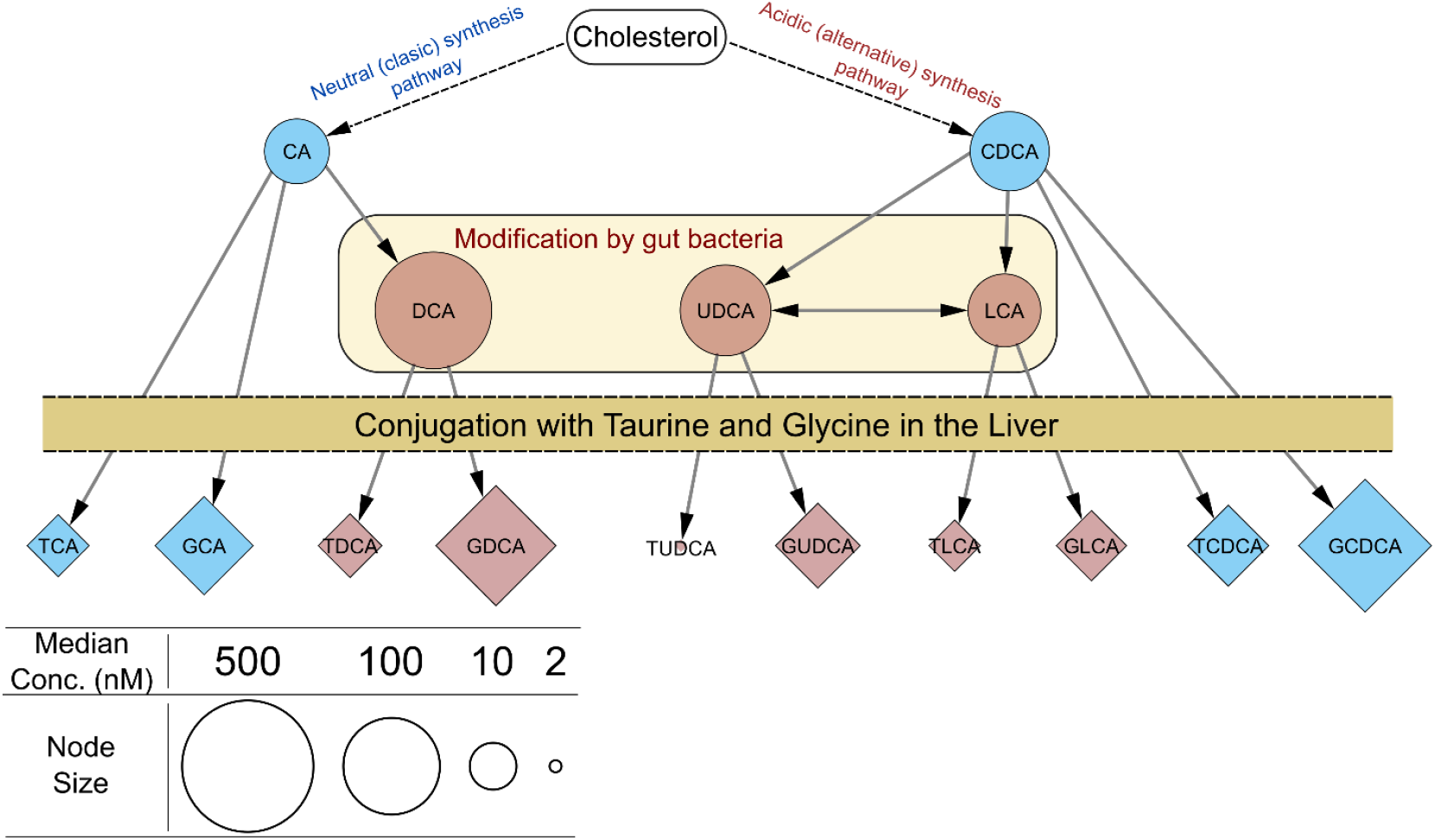
Bile acids biosynthesis pathway. Node colors represent primary (blue) and secondary (dark red) bile acids. Node shape represent conjugated (diamond) and unconjugated (oval) bile acids. Node size represents median concentration in the experimental cohort. Cholesterol (top of the pathway) is converted to primary bile acids along two pathways, neutral and acidic [1]. Further, primary bile acids are secreted to gut, and portion of it being modify by the gut bacteria to secondary bile acids. Primary and secondary bile acids are reabsorbed into the blood stream and reenter the liver, where they are conjugated with amino acids glycine or taurine. Conjugated bile acids are then being secreted back to the gut along with primary bile acids. Gut bacteria can cleave conjugated amino acids off bile acids [2], and freed metabolites are recirculated. Therefore, plasma levels of conjugated bile acids can me a reflection of both liver and gut bacteria activity. Refference: 1.Pandak, W.M. and G. Kakiyama, The acidic pathway of bile acid synthesis: Not just an alternative pathway(). Liver Res, 2019. 3(2): p. 88-98; 2. Ridlon, J.M., et al., Consequences of bile salt biotransformations by intestinal bacteria. Gut Microbes, 2016. 7(1): p. 22-39.

**Figure S3.**
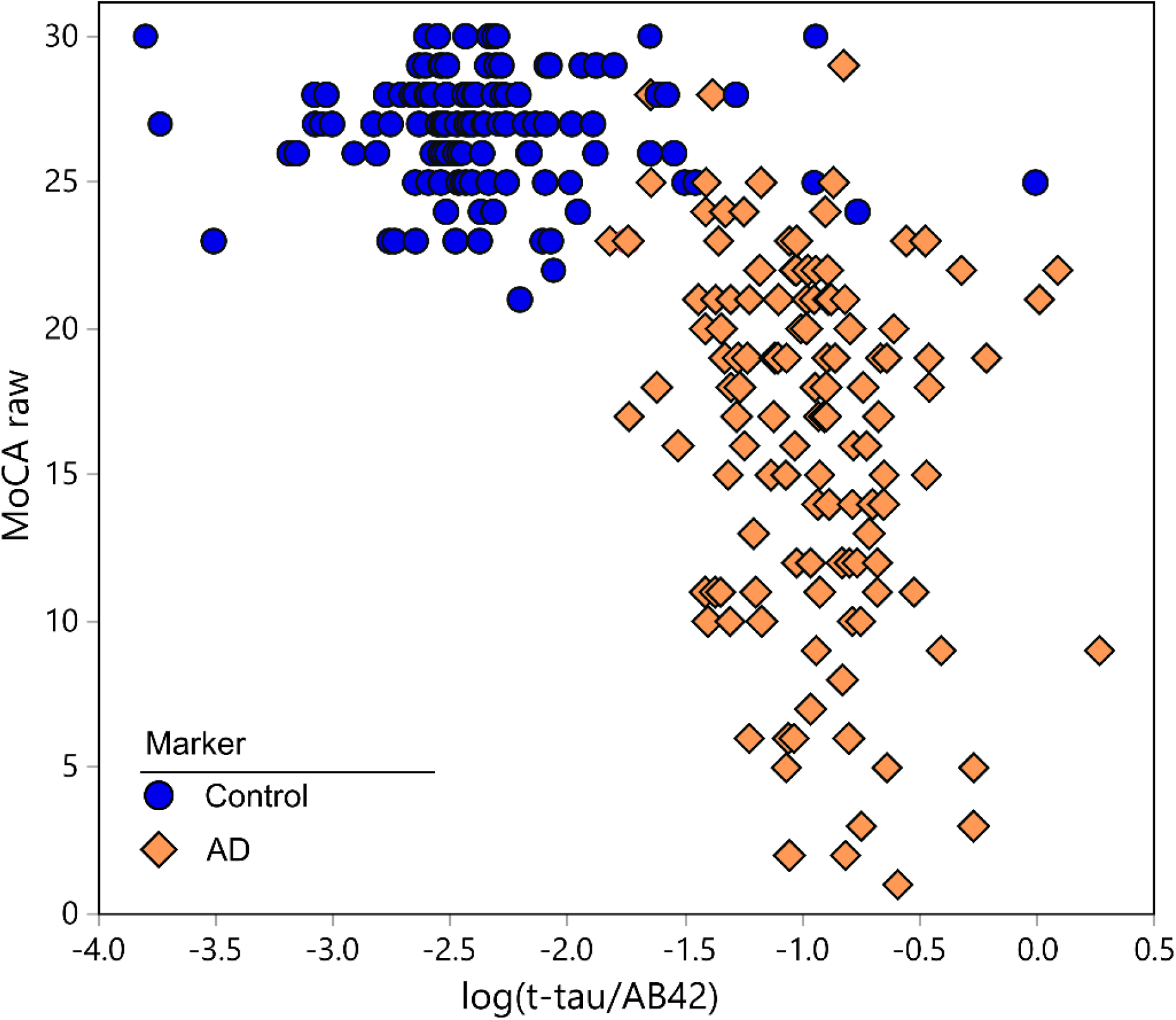
Control and AD group MoCA and log(t-Tau/Aβ42).

## References

1. Alkan, E., et al., Metabolic syndrome alters relationships between cardiometabolic variables, cognition and white matter hyperintensity load. Sci Rep, 2019. 9(1): p. 4356.

2. Bosia, M., et al., Improving Cognition to Increase Treatment Efficacy in Schizophrenia: Effects of Metabolic Syndrome on Cognitive Remediation’s Outcome. Front Psychiatry, 2018. 9: p. 647.

3. Monthe-Dreze, C., et al., Maternal obesity and offspring cognition: the role of inflammation. Pediatr Res, 2019. 85(6): p. 799–806.

4. Gabbs, M., et al., Advances in Our Understanding of Oxylipins Derived from Dietary PUFAs. Adv Nutr, 2015. 6(5): p. 513–40.

5. Nayeem, M.A., Role of oxylipins in cardiovascular diseases. Acta Pharmacol Sin, 2018. 39(7): p. 1142–1154.

6. Huang, C.C., et al., Association of Arachidonic Acid-derived Lipid Mediators with Subsequent Onset of Acute Myocardial Infarction in Patients with Coronary Artery Disease. Sci Rep, 2020. 10(1): p. 8105.

7. Ma, H. and M.E. Patti, Bile acids, obesity, and the metabolic syndrome. Best Pract Res Clin Gastroenterol, 2014. 28(4): p. 573–83.

8. Bellocchio, L., et al., The endocannabinoid system and energy metabolism. J Neuroendocrinol, 2008. 20(6): p. 850–7.

9. Zou, S. and U. Kumar, Cannabinoid Receptors and the Endocannabinoid System: Signaling and Function in the Central Nervous System. Int J Mol Sci, 2018. 19(3).

10. Chiurchiu, V., et al., The endocannabinoid system and its therapeutic exploitation in multiple sclerosis: Clues for other neuroinflammatory diseases. Prog Neurobiol, 2018. 160: p. 82–100.

11. Chiang, J.Y., Bile acid metabolism and signaling. Compr Physiol, 2013. 3(3): p. 1191–212.

12. Guo, C., W.D. Chen, and Y.D. Wang, TGR5, Not Only a Metabolic Regulator. Front Physiol, 2016. 7: p. 646.

13. Bazan, N.G., V. Colangelo, and W.J. Lukiw, Prostaglandins and other lipid mediators in Alzheimer’s disease. Prostaglandins Other Lipid Mediat, 2002. 68-69: p. 197–210.

14. Kao, Y.C., et al., Lipids and Alzheimer’s Disease. Int J Mol Sci, 2020. 21(4).

15. Miyazawa, K., et al., Alzheimer’s Disease and Specialized Pro-Resolving Lipid Mediators: Do MaR1, RvD1, and NPD1 Show Promise for Prevention and Treatment? Int J Mol Sci, 2020. 21(16).

16. MahmoudianDehkordi, S., et al., Altered bile acid profile associates with cognitive impairment in Alzheimer’s disease-An emerging role for gut microbiome. Alzheimers Dement, 2019. 15(1): p. 76–92.

17. Baloni, P., et al., Metabolic Network Analysis Reveals Altered Bile Acid Synthesis and Metabolism in Alzheimer’s Disease. Cell Rep Med, 2020. 1(8): p. 100138.

18. Ackerman, H.D. and G.S. Gerhard, Bile Acids in Neurodegenerative Disorders. Front Aging Neurosci, 2016. 8: p. 263.

19. Borkowski, K.e.a., Serum metabolomic biomarkers of perceptual speed in cognitively normal and mildly impaired subjects with fasting state stratification. https://doi.org/10.1101/2020.09.03.282343, 2020.

20. Goetz, M.E., et al., Rationale and Design of the Emory Healthy Aging and Emory Healthy Brain Studies. Neuroepidemiology, 2019. 53(3-4): p. 187–200.

21. Shaw, L.M., et al., Cerebrospinal fluid biomarker signature in Alzheimer’s disease neuroimaging initiative subjects. Ann Neurol, 2009. 65(4): p. 403–13.

22. Hulstaert, F., et al., Improved discrimination of AD patients using beta-amyloid(1-42) and tau levels in CSF. Neurology, 1999. 52(8): p. 1555–62.

23. Pedersen, T.L., I.J. Gray, and J.W. Newman, Plasma and serum oxylipin, endocannabinoid, bile acid, steroid, fatty acid and nonsteroidal anti-inflammatory drug quantification in a 96-well plate format. Anal Chim Acta, 2021. 1143: p. 189–200.

24. Agrawal, K., et al., Sweat lipid mediator profiling: a noninvasive approach for cutaneous research. J Lipid Res, 2017. 58(1): p. 188–195.

25. Saito, K., et al., Characterization of Postprandial Effects on CSF Metabolomics: A Pilot Study with Parallel Comparison to Plasma. Metabolites, 2020. 10(5).

26. Benjamini, Y. and Y. Hochberg, Controlling the False Discovery Rate: A Practical and Powerful Approach to Multiple Testing. Journal of the Royal Statistical Society. Series B (Methodological), 1995. 57(1): p. 289–300.

27. Lee, C.R., et al., Genetic variation in soluble epoxide hydrolase (EPHX2) and risk of coronary heart disease: The Atherosclerosis Risk in Communities (ARIC) study. Hum Mol Genet, 2006. 15(10): p. 1640–9.

28. Jahn, D. and A. Geier, Bile acids in nonalcoholic steatohepatitis: Pathophysiological driving force or innocent bystanders? Hepatology, 2018. 67(2): p. 464–466.

29. Grant, S.M. and S. DeMorrow, Bile Acid Signaling in Neurodegenerative and Neurological Disorders. Int J Mol Sci, 2020. 21(17).

30. Neu, S.C., et al., Apolipoprotein E Genotype and Sex Risk Factors for Alzheimer Disease: A Meta-analysis. JAMA Neurol, 2017. 74(10): p. 1178–1189.

31. Ghosh A, e.a., Epoxy fatty acid dysregulation and neuroinflammation in Alzheimer’s disease is resolved by a soluble epoxide hydrolase inhibitor. BioRxiv doi: https://doi.org/10.1101/2020.06.30.180984.

32. Kodani, S.D. and C. Morisseau, Role of epoxy-fatty acids and epoxide hydrolases in the pathology of neuro-inflammation. Biochimie, 2019. 159: p. 59–65.

33. Deng, Y., K.N. Theken, and C.R. Lee, Cytochrome P450 epoxygenases, soluble epoxide hydrolase, and the regulation of cardiovascular inflammation. J Mol Cell Cardiol, 2010. 48(2): p. 331–41.

34. Ulu, A., et al., Anti-inflammatory effects of omega-3 polyunsaturated fatty acids and soluble epoxide hydrolase inhibitors in angiotensin-II-dependent hypertension. J Cardiovasc Pharmacol, 2013. 62(3): p. 285–97.

35. Imig, J.D. and B.D. Hammock, Soluble epoxide hydrolase as a therapeutic target for cardiovascular diseases. Nat Rev Drug Discov, 2009. 8(10): p. 794–805.

36. Wagner, K.M., et al., Soluble epoxide hydrolase as a therapeutic target for pain, inflammatory and neurodegenerative diseases. Pharmacol Ther, 2017. 180: p. 62–76.

37. Firoz, C.K., et al., An overview on the correlation of neurological disorders with cardiovascular disease. Saudi J Biol Sci, 2015. 22(1): p. 19–23.

38. Klett, E.L., et al., Diminished acyl-CoA synthetase isoform 4 activity in INS 832/13 cells reduces cellular epoxyeicosatrienoic acid levels and results in impaired glucose-stimulated insulin secretion. J Biol Chem, 2013. 288(30): p. 21618–29.

39. Greene, J.F., et al., Metabolism of monoepoxides of methyl linoleate: bioactivation and detoxification. Arch Biochem Biophys, 2000. 376(2): p. 420–32.

40. Hennebelle, M., et al., Linoleic acid-derived metabolites constitute the majority of oxylipins in the rat pup brain and stimulate axonal growth in primary rat cortical neuron-glia co-cultures in a sex-dependent manner. J Neurochem, 2020. 152(2): p. 195–207.

41. Takahashi, H., et al., [Leukotoxin, 9,10-epoxy-12-octadecenoate, causes vasodilation in isolated pulmonary artery rings preconstricted with endothelin 1]. Nihon Kyobu Shikkan Gakkai Zasshi, 1992. 30(3): p. 418–24.

42. Hildreth, K., et al., Cytochrome P450-derived linoleic acid metabolites EpOMEs and DiHOMEs: a review of recent studies. J Nutr Biochem, 2020. 86: p. 108484.

43. Moghaddam, M.F., et al., Bioactivation of leukotoxins to their toxic diols by epoxide hydrolase. Nat Med, 1997. 3(5): p. 562–6.

44. Green, D., et al., Central activation of TRPV1 and TRPA1 by novel endogenous agonists contributes to mechanical allodynia and thermal hyperalgesia after burn injury. Mol Pain, 2016. 12.

45. Balleza-Tapia, H., et al., TrpV1 receptor activation rescues neuronal function and network gamma oscillations from Abeta-induced impairment in mouse hippocampus in vitro. Elife, 2018. 7.

46. Du, Y., et al., TRPV1 activation alleviates cognitive and synaptic plasticity impairments through inhibiting AMPAR endocytosis in APP23/PS45 mouse model of Alzheimer’s disease. Aging Cell, 2020. 19(3): p. e13113.

47. Raboune, S., et al., Novel endogenous N-acyl amides activate TRPV1-4 receptors, BV-2 microglia, and are regulated in brain in an acute model of inflammation. Front Cell Neurosci, 2014. 8: p. 195.

48. Borkowski, K., et al., Walnuts change lipoprotein composition suppressing TNFalpha-stimulated cytokine production by diabetic adipocyte. J Nutr Biochem, 2019. 68: p. 51–58.

49. Mahley, R.W., Central Nervous System Lipoproteins: ApoE and Regulation of Cholesterol Metabolism. Arterioscler Thromb Vasc Biol, 2016. 36(7): p. 1305–15.

50. Yan, H.C., et al., CYP2J2 rs890293 polymorphism is associated with susceptibility to Alzheimer’s disease in the Chinese Han population. Neuroscience Letters, 2015. 593: p. 56–60.

51. Srivastava, P.K., et al., Polymorphisms in human soluble epoxide hydrolase: effects on enzyme activity, enzyme stability, and quaternary structure. Arch Biochem Biophys, 2004. 427(2): p. 164–9.

52. Koerner, I.P., et al., Polymorphisms in the human soluble epoxide hydrolase gene EPHX2 linked to neuronal survival after ischemic injury. J Neurosci, 2007. 27(17): p. 4642–9.

53. Nelson, J.W., et al., Role of soluble epoxide hydrolase in age-related vascular cognitive decline. Prostaglandins Other Lipid Mediat, 2014. 113-115: p. 30–7.

54. Wingo, A.P., et al., Integrating human brain proteomes with genome-wide association data implicates new proteins in Alzheimer’s disease pathogenesis. Nat Genet, 2021. 53(2): p. 143–146.

55. Turcotte, C., et al., Regulation of inflammation by cannabinoids, the endocannabinoids 2-arachidonoyl-glycerol and arachidonoyl-ethanolamide, and their metabolites. J Leukoc Biol, 2015. 97(6): p. 1049–70.

56. Petrosino, S. and V. Di Marzo, The pharmacology of palmitoylethanolamide and first data on the therapeutic efficacy of some of its new formulations. Br J Pharmacol, 2017. 174(11): p. 1349–1365.

57. Saito, V.M., R.M. Rezende, and A.L. Teixeira, Cannabinoid modulation of neuroinflammatory disorders. Curr Neuropharmacol, 2012. 10(2): p. 159–66.

58. Bradshaw, H.B. and J.M. Walker, The expanding field of cannabimimetic and related lipid mediators. Br J Pharmacol, 2005. 144(4): p. 459–65.

59. Bradshaw, H.B., S. Raboune, and J.L. Hollis, Opportunistic activation of TRP receptors by endogenous lipids: exploiting lipidomics to understand TRP receptor cellular communication. Life Sci, 2013. 92(8-9): p. 404–9.

60. Li, K., et al., Interactions between sleep disturbances and Alzheimer’s disease on brain function: a preliminary study combining the static and dynamic functional MRI. Sci Rep, 2019. 9(1): p. 19064.

61. Koethe, D., et al., Sleep deprivation increases oleoylethanolamide in human cerebrospinal fluid. J Neural Transm (Vienna), 2009. 116(3): p. 301–5.

62. Giordano, C., et al., n-3 Polyunsaturated Fatty Acid Amides: New Avenues in the Prevention and Treatment of Breast Cancer. Int J Mol Sci, 2020. 21(7).

63. Villapol, S., Roles of Peroxisome Proliferator-Activated Receptor Gamma on Brain and Peripheral Inflammation. Cell Mol Neurobiol, 2018. 38(1): p. 121–132.

64. Berry, A.J., et al., Endocannabinoid system alterations in Alzheimer’s disease: A systematic review of human studies. Brain Res, 2020. 1749: p. 147135.

65. Aso, E. and I. Ferrer, Cannabinoids for treatment of Alzheimer’s disease: moving toward the clinic. Front Pharmacol, 2014. 5: p. 37.

66. Pedersen, T.L. and J.W. Newman, Establishing and Performing Targeted Multi-residue Analysis for Lipid Mediators and Fatty Acids in Small Clinical Plasma Samples. Methods Mol Biol, 2018. 1730: p. 175–212.

