## Supplementary material for "Association of plasma and CSF cytochrome P450, soluble epoxide hydrolase and ethanolamides metabolism with Alzheimer’s disease": Table S1

**Table S1: Cohort Characteristics**

| | CSF | | Plasma | |
|---|---|---|---|---|
| | **AD (n=150)** | **Control (n=139)** | **AD (n=148)** | **Control (n=133)** |
| Male:Females | 47%:53% | 28%:72% | 47%:53% | 27%:73% |
| Age[a] | 68.2 ± 1.34 | 65.2 ± 1.38 | 68 ± 1.36 | 65.6 ± 1.53 |
| AB42[a] | 300 ± 17.4 | 536 ± 25.8 | 300 ± 17.4 | 530 ± 25.8 |
| MoCA[a] | 16.4 ± 1.02 | 26.5 ± 0.523 | 16.4 ± 1.04 | 26.5 ± 0.579 |
| pTau[a] | 62.8 ± 3.88 | 31.6 ± 2.38 | 62.8 ± 3.93 | 30.7 ± 2.44 |
| tTau[a] | 115 ± 7.58 | 54.1 ± 4.09 | 115 ± 7.71 | 53.7 ± 4.6 |
| ApoE genotype % | | | | |
| E2/E3 | 1% | 17% | 1% | 17% |
| E2/E4 | 2% | 5% | 2% | 4% |
| E3/E3 | 27% | 53% | 27% | 55% |
| E3/E4 | 51% | 22% | 50% | 21% |
| E4/E4 | 19% | 3% | 20% | 3% |
| Race % | | | | |
| Native americans | 1% | - | 1% | - |
| Asian | 1% | 1% | 1% | - |
| Black or African American | 7% | 14% | 7% | 14% |
| Caucasian or White | 92% | 86% | 92% | 86% |

*a* – Results are means ± 95% confidence intervals
