## Supplementary material for "Association of plasma and CSF cytochrome P450, soluble epoxide hydrolase and ethanolamides metabolism with Alzheimer’s disease": Table S2

**Table S2.** T-test of predicted fasted vs predicted non-fasted in AD group for plasma and CSF metabolites.

| CSF | | | | |
|---|---|---|---|---|
| **Metabolite** | **P value** | **Fold change** | **Mean (nM) [95%CI]** | |
| | | | **Fasted** | **Non-fasted** |
| PGF2a | 0.0251 | 1.25 | 0.0274 [0.0235-0.032] | 0.0342 [0.0301-0.0388] |
| GCA/GCDCA | 0.0263 | 1.18 | 1.12 [0.993-1.27] | 1.32 [1.15-1.52] |
| 12_13-DiHOME | 0.03 | 1.27 | 0.0305 [0.0268-0.0347] | 0.0387 [0.0328-0.0457] |
| UDCA/CDCA | 0.0472 | 0.585 | 0.978 [0.683-1.4] | 0.572 [0.441-0.744] |
| 12,13-DiHOME/EpOME | 0.0626 | 1.24 | 0.0918 [0.077-0.109] | 0.114 [0.0939-0.137] |
| OEA | 0.07 | 1.13 | 0.082 [0.0757-0.0888] | 0.0923 [0.0832-0.102] |
| DCA/(LCA+UDCA) | 0.0737 | 1.52 | 1.83 [1.28-2.62] | 2.79 [2.13-3.65] |
| TCA | 0.0742 | 1.38 | 0.407 [0.295-0.561] | 0.563 [0.447-0.711] |
| 15_16-DiHODE | 0.0875 | 1.16 | 0.0669 [0.0567-0.0789] | 0.0775 [0.0666-0.0902] |
| GCA | 0.0925 | 1.25 | 0.887 [0.712-1.11] | 1.11 [0.897-1.37] |
| LA_RelAb | 0.108 | 0.915 | 0.248 [0.228-0.269] | 0.227 [0.212-0.244] |
| GLCA/CDCA | 0.114 | 0.674 | 0.0362 [0.0263-0.0497] | 0.0244 [0.0187-0.0318] |
| Sum(DiHOME)/Sum(EpOME) | 0.121 | 1.2 | 0.0794 [0.066-0.0955] | 0.0954 [0.0788-0.115] |
| 9_10-DiHOME | 0.122 | 1.17 | 0.0197 [0.017-0.0228] | 0.023 [0.0194-0.0273] |
| GDCA/GLCA | 0.14 | 1.34 | 10.1 [7.44-13.6] | 13.5 [10.7-17.2] |
| UDCA | 0.153 | 0.693 | 1.12 [0.767-1.62] | 0.776 [0.593-1.01] |
| (GDCA+TDCA)/(TLCA+GLCA) | 0.154 | 1.33 | 11.4 [8.4-15.5] | 15.2 [12-19.4] |
| GCDCA/GLCA | 0.169 | 1.32 | 19.2 [14.8-24.8] | 25.4 [20.6-31.2] |
| TUDCA/UDCA | 0.189 | 1.46 | 0.00304 [0.00188-0.00489] | 0.00445 [0.0031-0.00639] |
| GLCA | 0.226 | 0.799 | 0.0413 [0.0332-0.0512] | 0.033 [0.0269-0.0406] |
| EPA + DHA diols | 0.236 | 0.866 | 0.157 [0.137-0.181] | 0.136 [0.119-0.155] |
| 17_18-DiHETE | 0.268 | 0.768 | 0.104 [0.0877-0.124] | 0.0799 [0.063-0.101] |
| 9,10-DiHOME/EpOME | 0.269 | 1.14 | 0.0641 [0.0517-0.0794] | 0.0728 [0.0588-0.09] |
| GCA/GDCA | 0.27 | 1.16 | 2.14 [1.66-2.75] | 2.48 [1.96-3.14] |
| 13-HOTE | 0.273 | 1.08 | 0.0478 [0.0419-0.0546] | 0.0517 [0.047-0.0569] |
| GUDCA/UDCA | 0.291 | 1.37 | 0.0921 [0.0617-0.137] | 0.126 [0.0904-0.176] |
| (TUDAC+GUDCA)/UDCA | 0.295 | 1.36 | 0.0976 [0.0656-0.145] | 0.133 [0.0954-0.184] |
| ALA_RelAb | 0.297 | 0.949 | 0.272 [0.253-0.293] | 0.258 [0.24-0.276] |
| (GDCA+GLCA)/(TDCA) | 0.31 | 0.905 | 11.6 [9.19-14.6] | 10.5 [9.13-12.1] |
| 19_20-DiHDoPE | 0.333 | 0.918 | 0.0183 [0.0157-0.0213] | 0.0168 [0.0151-0.0188] |
| AA_RelAb | 0.354 | 0.941 | 0.256 [0.234-0.281] | 0.241 [0.223-0.261] |
| CDCA | 0.375 | 1.19 | 1.14 [0.886-1.47] | 1.36 [1.11-1.65] |
| GCDCA/CDCA | 0.376 | 0.892 | 0.693 [0.532-0.903] | 0.618 [0.525-0.727] |
| TCDCA | 0.387 | 1.11 | 0.0656 [0.051-0.0845] | 0.0726 [0.0583-0.0903] |
| (TCDCA+GCDCA)/CDCA | 0.396 | 0.889 | 0.767 [0.587-1] | 0.682 [0.578-0.805] |
| a-MCA | 0.4 | 1.22 | 0.0316 [0.0212-0.047] | 0.0387 [0.0291-0.0515] |
| 20-HETE | 0.41 | 1.07 | 0.121 [0.107-0.136] | 0.13 [0.118-0.143] |
| T-a-MCA | 0.415 | 1.06 | 0.0294 [0.0229-0.0378] | 0.0311 [0.0243-0.0399] |
| 13-HODE | 0.416 | 1.03 | 1.56 [1.51-1.61] | 1.61 [1.55-1.68] |
| EPA_RelAb | 0.418 | 0.915 | 0.211 [0.179-0.25] | 0.193 [0.168-0.222] |

| | | | | |
|---|---|---|---|---|
| TCDCA/GCDCA | 0.421 | 1.04 | 0.083 [0.0683-0.101] | 0.0867 [0.0742-0.101] |
| GDCA | 0.427 | 1.08 | 0.415 [0.325-0.531] | 0.447 [0.364-0.549] |
| 14,15/11,12-DiHETrE | 0.428 | 1.03 | 2.74 [2.59-2.9] | 2.82 [2.67-2.98] |
| 9-HOTE | 0.428 | 1.05 | 0.0119 [0.0106-0.0133] | 0.0125 [0.0112-0.0139] |
| 14_15-DiHETE | 0.431 | 0.874 | 0.0247 [0.0196-0.0312] | 0.0216 [0.0168-0.0279] |
| TDCA | 0.432 | 1.14 | 0.0422 [0.0303-0.0588] | 0.0479 [0.0377-0.0609] |
| TEST | 0.434 | 1.11 | 0.0426 [0.0301-0.0602] | 0.0472 [0.0358-0.0623] |
| 17OH-PROG | 0.436 | 1.1 | 0.0436 [0.0381-0.0498] | 0.0479 [0.0424-0.0541] |
| 11_12-DiHETrE | 0.45 | 0.954 | 0.0175 [0.0154-0.0199] | 0.0167 [0.0151-0.0184] |
| w-MCA/UDCA | 0.456 | 1.34 | 0.107 [0.071-0.162] | 0.143 [0.0968-0.211] |
| w-MCA/T-a-MCA | 0.528 | 0.875 | 4.07 [2.54-6.53] | 3.56 [2.23-5.67] |
| TDCA/GDCA | 0.57 | 1.05 | 0.102 [0.0823-0.126] | 0.107 [0.0937-0.123] |
| (TDCA+TCDCA)/(GDCA+GCDCA) | 0.587 | 1.03 | 0.0934 [0.0775-0.113] | 0.0958 [0.0837-0.11] |
| LEA | 0.59 | 1 | 0.255 [0.237-0.274] | 0.256 [0.245-0.268] |
| DHEA | 0.592 | 0.939 | 0.0163 [0.0141-0.0188] | 0.0153 [0.0138-0.0171] |
| DCA | 0.609 | 1.06 | 2.04 [1.58-2.64] | 2.16 [1.7-2.75] |
| (GDCA+TDCA)/(TUDCA+GUDCA) | 0.62 | 1.13 | 4.33 [2.96-6.32] | 4.89 [3.65-6.54] |
| TDCA/DCA | 0.626 | 1.07 | 0.0207 [0.0144-0.0296] | 0.0222 [0.0174-0.0283] |
| TDCA/DCA 2 | 0.626 | 1.07 | 0.0207 [0.0144-0.0296] | 0.0222 [0.0174-0.0283] |
| w-MCA | 0.634 | 0.925 | 0.12 [0.0816-0.176] | 0.111 [0.0778-0.158] |
| CRTN | 0.701 | 1.03 | 4.26 [3.89-4.66] | 4.37 [4.1-4.66] |
| GCDCA/GDCA | 0.744 | 0.984 | 1.9 [1.52-2.39] | 1.87 [1.51-2.32] |
| 11-Deoxy-CTRL | 0.746 | 0.948 | 0.0541 [0.0474-0.0617] | 0.0513 [0.0454-0.058] |
| GUDCA | 0.769 | 0.949 | 0.103 [0.0766-0.138] | 0.0977 [0.0771-0.124] |
| TUDCA | 0.769 | 1.02 | 0.00339 [0.0025-0.00459] | 0.00345 [0.00271-0.0044] |
| F2-IsoP | 0.79 | 0.97 | 0.301 [0.28-0.323] | 0.292 [0.272-0.314] |
| CRTL | 0.855 | 1 | 14.4 [13.1-15.9] | 14.4 [13.5-15.4] |
| 12(13)-EpOME | 0.863 | 1.03 | 0.332 [0.285-0.387] | 0.341 [0.298-0.391] |
| TCDCA/CDCA | 0.887 | 0.932 | 0.0575 [0.0402-0.0822] | 0.0536 [0.0412-0.0696] |
| GCDCA | 0.889 | 1.06 | 0.79 [0.658-0.949] | 0.837 [0.723-0.97] |
| 14_15-DiHETrE | 0.901 | 0.981 | 0.0479 [0.043-0.0535] | 0.047 [0.0435-0.0508] |
| DHA_RelAb | 0.925 | 0.988 | 0.256 [0.224-0.293] | 0.253 [0.224-0.286] |
| 9-HODE | 0.931 | 1.02 | 0.532 [0.511-0.553] | 0.544 [0.518-0.571] |
| GDCA/DCA | 0.934 | 1.02 | 0.203 [0.166-0.249] | 0.207 [0.179-0.239] |
| (TDCA+GDCA)/DCA | 0.937 | 1.01 | 0.231 [0.186-0.286] | 0.233 [0.2-0.271] |
| CRCTN | 0.954 | 0.998 | 0.414 [0.35-0.49] | 0.413 [0.361-0.472] |
| 9(10)-EpOME | 0.975 | 1.03 | 0.307 [0.257-0.366] | 0.317 [0.268-0.374] |
| T-a-MCA/CDCA | 0.995 | 0.891 | 0.0258 [0.0181-0.0368] | 0.023 [0.0164-0.0322] |
| Plasma | | | | |
| 12(13)-EpOME | 0.0001 | 2.79 | 2.41 [2.07-2.8] | 6.73 [5.54-8.17] |
| 12_13-DiHODE | 0.0001 | 2.52 | 0.209 [0.161-0.271] | 0.527 [0.448-0.62] |
| 12_13-DiHOME | 0.0001 | 2.29 | 3.86 [3.38-4.41] | 8.83 [7.62-10.2] |
| 13-HOTE | 0.0001 | 2.26 | 0.624 [0.483-0.806] | 1.41 [1.17-1.7] |
| 15(16)-EpODE | 0.0001 | 2.17 | 2.42 [2.06-2.84] | 5.24 [4.33-6.35] |
| 15_16-DiHODE | 0.0001 | 2.28 | 10.9 [9.47-12.5] | 24.9 [21.5-28.9] |
| 9(10)-EpOME | 0.0001 | 1.92 | 0.372 [0.305-0.454] | 0.713 [0.574-0.885] |

| | | | | |
|---|---|---|---|---|
| 9_10-DiHODE | 0.0001 | 4.33 | 0.183 [0.144-0.233] | 0.792 [0.629-0.997] |
| 9_10-DiHOME | 0.0001 | 2.93 | 3.25 [2.94-3.59] | 9.53 [8.04-11.3] |
| 9_10-e-DiHO | 0.0001 | 1.56 | 3.05 [2.68-3.46] | 4.75 [4.07-5.54] |
| 9-HOTE | 0.0001 | 1.72 | 0.446 [0.39-0.511] | 0.769 [0.652-0.906] |
| AA_screen | 0.0001 | 0.645 | 0.155 [0.139-0.172] | 0.1 [0.0895-0.113] |
| AEA | 0.0001 | 0.691 | 1.49 [1.36-1.64] | 1.03 [0.92-1.15] |
| GCA | 0.0001 | 3.31 | 68.6 [51.6-91.1] | 227 [179-288] |
| GCDCA | 0.0001 | 2.97 | 367 [270-499] | 1090 [896-1330] |
| GDCA | 0.0001 | 2.83 | 186 [125-277] | 526 [399-694] |
| NA-Gly | 0.0001 | 0.619 | 0.72 [0.621-0.835] | 0.446 [0.389-0.512] |
| NO-Gly | 0.0001 | 0.53 | 4.7 [4.29-5.16] | 2.49 [2.18-2.85] |
| OEA | 0.0001 | 0.755 | 5.23 [4.79-5.7] | 3.95 [3.58-4.36] |
| TCA | 0.0001 | 3.56 | 11.7 [8.1-16.9] | 41.7 [32.3-53.9] |
| TCDCA | 0.0001 | 3.27 | 30.3 [21.6-42.7] | 99 [77.4-127] |
| TDCA | 0.0001 | 2.92 | 13.7 [8.95-21] | 40 [29.1-55.1] |
| POEA_Screen | 0.0002 | 0.603 | 0.119 [0.0985-0.144] | 0.0718 [0.0602-0.0856] |
| TLCA | 0.0002 | 2.3 | 7.56 [5.18-11.1] | 17.4 [13.2-23] |
| DHA_screen | 0.0003 | 0.652 | 0.149 [0.128-0.174] | 0.0971 [0.0838-0.113] |
| DHEA | 0.0003 | 0.726 | 1.18 [1.04-1.34] | 0.857 [0.78-0.941] |
| LA_screen | 0.0004 | 0.722 | 0.158 [0.138-0.18] | 0.114 [0.101-0.129] |
| 9_12_13-TriHOME | 0.0005 | 1.63 | 2.65 [2.2-3.18] | 4.32 [3.64-5.13] |
| GUDCA | 0.0006 | 2.31 | 46.3 [32.8-65.4] | 107 [80.8-142] |
| LEA | 0.0006 | 0.811 | 4.5 [4.13-4.91] | 3.65 [3.39-3.93] |
| ALA_screen | 0.0008 | 0.646 | 0.153 [0.128-0.184] | 0.0988 [0.0822-0.119] |
| GLCA | 0.0009 | 2.27 | 21.8 [14.8-32.1] | 49.5 [38.2-64.1] |
| 9-HODE | 0.0015 | 1.46 | 10.9 [9.66-12.2] | 15.9 [13.5-18.7] |
| DGLEA | 0.0016 | 0.726 | 0.175 [0.151-0.201] | 0.127 [0.11-0.147] |
| EPA_screen | 0.0017 | 0.625 | 0.13 [0.108-0.156] | 0.0813 [0.0672-0.0983] |
| 13-HODE | 0.0023 | 1.45 | 14.3 [12.7-16.1] | 20.7 [17.5-24.5] |
| GHDCA | 0.0023 | 1.88 | 0.8 [0.611-1.05] | 1.5 [1.24-1.82] |
| T-a-MCA | 0.0037 | 1.95 | 2.62 [1.9-3.63] | 5.1 [3.86-6.73] |
| 5-HETE | 0.0056 | 0.755 | 3.79 [3.35-4.29] | 2.86 [2.52-3.23] |
| DEA | 0.0072 | 0.788 | 0.378 [0.325-0.44] | 0.298 [0.266-0.334] |
| TUDCA | 0.0084 | 1.69 | 1.92 [1.54-2.4] | 3.24 [2.56-4.09] |
| 12-HETE | 0.0137 | 0.786 | 2.81 [2.46-3.21] | 2.21 [1.95-2.51] |
| 15-HETE | 0.0202 | 0.824 | 3.13 [2.81-3.49] | 2.58 [2.28-2.92] |
| Ibuprofen | 0.0266 | 0.427 | 41 [21.4-78.4] | 17.5 [10.9-27.9] |
| w-MCA | 0.0392 | 1.78 | 7.68 [5.53-10.7] | 13.7 [10.2-18.4] |
| CDCA | 0.0401 | 1.9 | 46.7 [32.6-66.9] | 88.5 [61.6-127] |
| 1-LG | 0.0436 | 1.23 | 175 [155-197] | 215 [192-241] |
| aLEA | 0.0483 | 0.837 | 0.184 [0.164-0.207] | 0.154 [0.138-0.172] |
| DCA | 0.0893 | 1.55 | 177 [120-259] | 274 [206-366] |
| TEST | 0.0953 | 0.924 | 2.5 [1.63-3.85] | 2.31 [1.6-3.34] |
| 11_12-DiHETrE | 0.0989 | 0.877 | 0.81 [0.735-0.894] | 0.71 [0.64-0.788] |
| 11-HETE | 0.1022 | 0.845 | 1.61 [1.4-1.86] | 1.36 [1.19-1.56] |
| 9(10)-EpODE | 0.1086 | 1.32 | 0.145 [0.112-0.187] | 0.192 [0.146-0.253] |
| 9-HETE | 0.1162 | 0.803 | 1.32 [1.12-1.57] | 1.06 [0.894-1.27] |

| | | | | |
|---|---|---|---|---|
| 2-LG | 0.1187 | 1.19 | 46.4 [41-52.5] | 55 [49.4-61.3] |
| 13-KODE | 0.1265 | 0.732 | 1.42 [1.16-1.74] | 1.04 [0.809-1.33] |
| 11(12)-EpETrE | 0.1495 | 0.697 | 0.0755 [0.0608-0.0938] | 0.0526 [0.0415-0.0665] |
| 14(15)-EpETrE | 0.1565 | 1.07 | 0.106 [0.0826-0.136] | 0.113 [0.0936-0.137] |
| PGF2a | 0.1565 | 1.35 | 0.416 [0.313-0.554] | 0.56 [0.473-0.664] |
| 14-HDoHE | 0.1646 | 0.804 | 1.58 [1.19-2.11] | 1.27 [0.96-1.67] |
| CA | 0.1729 | 1.66 | 19.3 [13.6-27.4] | 32 [21.7-47.2] |
| Progesterone | 0.1736 | 1.26 | 0.37 [0.317-0.432] | 0.466 [0.39-0.558] |
| EPEA_Screen | 0.1844 | 0.808 | 0.0873 [0.0704-0.108] | 0.0705 [0.0581-0.0855] |
| 14_15-DiHETrE | 0.2208 | 0.922 | 1 [0.912-1.1] | 0.922 [0.83-1.02] |
| 8_9-DiHETrE | 0.249 | 0.742 | 0.365 [0.279-0.476] | 0.271 [0.215-0.342] |
| PGE2 | 0.2637 | 1.22 | 0.16 [0.13-0.197] | 0.195 [0.16-0.238] |
| 5_6-DiHETrE | 0.2763 | 0.875 | 0.51 [0.449-0.581] | 0.446 [0.384-0.518] |
| Naproxen | 0.3047 | 0.565 | 191 [57.1-637] | 108 [40-294] |
| 5-HEPE | 0.3191 | 0.82 | 0.572 [0.463-0.706] | 0.469 [0.387-0.568] |
| b-MCA | 0.3361 | 0.813 | 2.41 [1.79-3.24] | 1.96 [1.55-2.49] |
| 12-HEPE | 0.4055 | 0.9 | 0.311 [0.252-0.383] | 0.28 [0.236-0.332] |
| Acetaminophen | 0.4853 | 1.11 | 5.52 [2.81-10.8] | 6.14 [3.36-11.2] |
| 1-AG | 0.5174 | 1.07 | 19 [16.9-21.4] | 20.4 [18.2-22.8] |
| UDCA | 0.5175 | 0.877 | 91.6 [62.8-134] | 80.3 [59.2-109] |
| LCA | 0.5555 | 1.12 | 30 [23.7-38] | 33.6 [27.2-41.5] |
| 9-HEPE | 0.5787 | 1.02 | 0.158 [0.111-0.225] | 0.161 [0.132-0.196] |
| 1-OG | 0.595 | 1.09 | 282 [252-316] | 308 [276-344] |
| 2-AG | 0.6627 | 1.07 | 4.77 [4.24-5.37] | 5.12 [4.49-5.84] |
| CRCTN | 0.6773 | 1.09 | 0.743 [0.58-0.953] | 0.808 [0.693-0.943] |
| CRTN | 0.6992 | 0.966 | 4.76 [3.71-6.11] | 4.6 [3.85-5.49] |
| 4-HDoHE | 0.717 | 0.951 | 0.649 [0.549-0.768] | 0.617 [0.506-0.751] |
| 17_18-DiHETE | 0.7633 | 1.01 | 5.23 [4.32-6.34] | 5.29 [4.49-6.23] |
| 15-HEPE | 0.7908 | 0.925 | 0.292 [0.239-0.356] | 0.27 [0.223-0.327] |
| 8-HETE | 0.8305 | 0.992 | 2.51 [2.16-2.93] | 2.49 [2.18-2.84] |
| TXB2 | 0.8681 | 1.03 | 0.231 [0.183-0.292] | 0.239 [0.193-0.298] |
| 19_20-DiHDoPE | 0.8845 | 0.985 | 2.03 [1.78-2.33] | 2 [1.8-2.23] |
| 17-OH PROG | 0.8963 | 1.03 | 0.951 [0.764-1.18] | 0.978 [0.805-1.19] |
| PGD2 | 0.9007 | 0.928 | 0.264 [0.176-0.397] | 0.245 [0.178-0.338] |
| CRTL | 0.9039 | 1.17 | 207 [158-272] | 242 [217-270] |
| 2-OG | 0.9312 | 1.05 | 34.1 [29.8-39.1] | 35.9 [31.6-40.7] |
| 5_15-DiHETE | 0.9689 | 1.1 | 0.105 [0.0756-0.146] | 0.115 [0.0904-0.146] |
| DHEAS | 0.9727 | 1.24 | 1220 [923-1610] | 1510 [1310-1740] |
| F2-IsoP | 0.9741 | 0.959 | 6.36 [5.33-7.59] | 6.1 [5.34-6.98] |
