## Supplementary material for "Association of plasma and CSF cytochrome P450, soluble epoxide hydrolase and ethanolamides metabolism with Alzheimer’s disease": Table S3

**Table S3.** The mean values and t-test and two-way ANOVA interaction p-values for all detected metabolites in plasma. P-values < 0.05 are colored red, p-values > 0.05 < 0.1 are colored orange.

| Metabolite information | | | | p-values | | | | Mean [95%CI] Uning subjects with p of fasted >60% | |
|---|---|---|---|---|---|---|---|---|---|
| | | | | Uning subjects with p of fasted >60% | | | **p-value** AD vs Control All subjects | | |
| Metabolite name | Chemical class | Enzyme | Units | **p-value** AD vs Control | **p-value** Gender x Disease interaction | **p-value** Race x Disease interaction | | Control mean [95%CI] | AD mean [95%CI] |
| Oxylipins, endocannabinoids, PUFAS and NSADs | | | | | | | | | |
| TXB2 | TX | COX1 | nM | 0.108 | 0.516 | 0.309 | 0.0435 | 0.327 [0.262 - 0.408] | 0.234 [0.178 - 0.307] |
| PGE2 | PGs | COX2 | nM | 0.112 | 0.275 | 0.236 | 0.0459 | 0.209 [0.182 - 0.238] | 0.16 [0.13 - 0.197] |
| PGD2 | PGs | COX2 | nM | 0.0346 | 0.327 | 0.0459 | 0.0037 | 0.355 [0.286 - 0.441] | 0.225 [0.142 - 0.355] |
| PGF2a | PGs | COX2 | nM | 0.0163 | 0.481 | 0.609 | 0.0632 | 0.65 [0.525 - 0.805] | 0.387 [0.284 - 0.527] |
| F2-IsoP | PGs | Auto-ox | nM | 0.0059 | 0.407 | 0.923 | 0.0001 | 8.22 [7.15 - 9.45] | 6.36 [5.33 - 7.59] |
| 5_15-DiHETE | Diol | LOX | nM | 0.272 | 0.278 | 0.381 | 0.383 | 0.0998 [0.0777 - 0.128] | 0.106 [0.0717 - 0.158] |
| 9_12_13-TriHOME | Triol | LOX | nM | 0.0006 | 0.828 | 0.165 | 0.0618 | 4.38 [3.75 - 5.1] | 2.65 [2.2 - 3.18] |
| 9_10-e-DiHO | vic-Diol | sEH | nM | 0.0236 | 0.905 | 0.605 | 0.986 | 3.97 [3.56 - 4.42] | 3.05 [2.68 - 3.46] |
| 12_13-DiHOME | vic-Diol | sEH | nM | 0.103 | 0.7 | 0.495 | 0.0761 | 4.71 [4.26 - 5.21] | 3.86 [3.38 - 4.41] |
| 9_10-DiHOME | vic-Diol | sEH | nM | 0.413 | 0.442 | 0.377 | 0.0001 | 3.16 [2.89 - 3.45] | 3.25 [2.94 - 3.59] |
| 15_16-DiHODE | vic-Diol | sEH | nM | 0.813 | 0.858 | 0.849 | 0.0001 | 11.6 [10.5 - 12.8] | 10.9 [9.47 - 12.5] |
| 12_13-DiHODE | vic-Diol | sEH | nM | 0.711 | 0.26 | 0.502 | 0.0001 | 0.218 [0.176 - 0.27] | 0.202 [0.151 - 0.27] |
| 9_10-DiHODE | vic-Diol | sEH | nM | 0.909 | 0.587 | 0.235 | 0.0001 | 0.192 [0.162 - 0.228] | 0.184 [0.141 - 0.24] |
| 14_15-DiHETrE | vic-Diol | sEH | nM | 0.0004 | 0.305 | 0.833 | 0.0017 | 0.833 [0.784 - 0.886] | 1 [0.912 - 1.1] |
| 11_12-DiHETrE | vic-Diol | sEH | nM | 0.0092 | 0.23 | 0.757 | 0.0538 | 0.709 [0.67 - 0.75] | 0.81 [0.735 - 0.894] |
| 8_9-DiHETrE | vic-Diol | sEH | nM | 0.205 | 0.762 | 0.97 | 0.87 | 0.266 [0.203 - 0.347] | 0.339 [0.24 - 0.478] |
| 5_6-DiHETrE | vic-Diol | sEH | nM | 0.47 | 0.507 | 0.653 | 0.0223 | 0.565 [0.513 - 0.622] | 0.51 [0.449 - 0.581] |
| 17_18-DiHETE | vic-Diol | sEH | nM | 0.0001 | 0.118 | 0.632 | 0.0001 | 1.73 [1.32 - 2.27] | 5.23 [4.32 - 6.34] |
| 19_20-DiHDoPE | vic-Diol | sEH | nM | 0.802 | 0.0983 | 0.938 | 0.623 | 2 [1.83 - 2.18] | 2.03 [1.78 - 2.33] |
| 13-HODE | R-OH | LOX | nM | 0.0793 | 0.305 | 0.96 | 0.76 | 16.8 [15.4 - 18.3] | 14.3 [12.7 - 16.1] |
| 9-HODE | R-OH | LOX | nM | 0.563 | 0.428 | 0.815 | 0.128 | 11.9 [10.9 - 13] | 10.9 [9.66 - 12.2] |
| 13-HOTE | R-OH | LOX | nM | 0.686 | 0.299 | 0.907 | 0.0082 | 0.709 [0.601 - 0.836] | 0.624 [0.483 - 0.806] |
| 9-HOTE | R-OH | LOX | nM | 0.863 | 0.976 | 0.0958 | 0.0077 | 0.479 [0.424 - 0.542] | 0.446 [0.39 - 0.511] |
| 15-HETE | R-OH | LOX | nM | 0.769 | 0.316 | 0.639 | 0.241 | 3.17 [2.82 - 3.55] | 3.13 [2.81 - 3.49] |
| 12-HETE | R-OH | LOX | nM | 0.224 | 0.751 | 0.0638 | 0.0005 | 3.35 [2.95 - 3.81] | 2.81 [2.46 - 3.21] |
| 11-HETE | R-OH | LOX | nM | 0.224 | 0.619 | 0.966 | 0.0021 | 2.01 [1.75 - 2.3] | 1.61 [1.4 - 1.86] |
| 9-HETE | R-OH | LOX | nM | 0.168 | 0.57 | 0.464 | 0.0039 | 1.73 [1.47 - 2.03] | 1.32 [1.12 - 1.57] |
| 8-HETE | R-OH | LOX | nM | 0.0032 | 0.984 | 0.659 | 0.0001 | 3.64 [3.18 - 4.17] | 2.51 [2.16 - 2.93] |
| 5-HETE | R-OH | LOX | nM | 0.0593 | 0.648 | 0.272 | 0.0001 | 5.02 [4.43 - 5.68] | 3.79 [3.35 - 4.29] |
| 15-HEPE | R-OH | LOX | nM | 0.116 | 0.235 | 0.979 | 0.0059 | 0.345 [0.282 - 0.422] | 0.294 [0.24 - 0.359] |
| 12-HEPE | R-OH | LOX | nM | 0.0073 | 0.598 | 0.792 | 0.0001 | 0.459 [0.39 - 0.539] | 0.313 [0.253 - 0.387] |
| 9-HEPE | R-OH | Auto-ox | nM | 0.0105 | 0.93 | 0.174 | 0.0001 | 0.319 [0.256 - 0.397] | 0.158 [0.103 - 0.241] |
| 5-HEPE | R-OH | LOX | nM | 0.0028 | 0.705 | 0.754 | 0.0001 | 0.908 [0.771 - 1.07] | 0.572 [0.463 - 0.706] |
| 14-HDoHE | R-OH | LOX | nM | 0.0171 | 0.324 | 0.574 | 0.0002 | 2.4 [2 - 2.88] | 1.61 [1.17 - 2.21] |
| 4-HDoHE | R-OH | LOX | nM | 0.0008 | 0.402 | 0.0713 | 0.0001 | 1.03 [0.886 - 1.21] | 0.651 [0.545 - 0.777] |
| 13-KODE | R=0 | ADH | nM | 0.349 | 0.614 | 0.261 | 0.0061 | 1.74 [1.43 - 2.12] | 1.46 [1.18 - 1.81] |
| 12(13)-EpOME | Epox | CYP | nM | 0.405 | 0.719 | 0.192 | 0.0001 | 2.28 [2.03 - 2.56] | 2.41 [2.07 - 2.8] |
| 9(10)-EpOME | Epox | CYP | nM | 0.026 | 0.9 | 0.452 | 0.0001 | 0.29 [0.244 - 0.346] | 0.369 [0.3 - 0.453] |
| 15(16)-EpODE | Epox | CYP | nM | 0.206 | 0.7 | 0.414 | 0.0001 | 2.14 [1.9 - 2.4] | 2.42 [2.06 - 2.84] |
| 9(10)-EpODE | Epox | CYP | nM | 0.456 | 0.283 | 0.16 | 0.63 | 0.174 [0.147 - 0.206] | 0.146 [0.11 - 0.193] |
| 14(15)-EpETrE | Epox | CYP | nM | 0.333 | 0.101 | 0.496 | 0.076 | 0.0851 [0.0634 - 0.114] | 0.1 [0.074 - 0.136] |
| 11(12)-EpETrE | Epox | CYP | nM | 0.3 | 0.349 | 0.259 | 0.616 | 0.0574 [0.0439 - 0.0751] | 0.0705 [0.0507 - 0.0979] |
| LA | PUFA | Diet | Rel Abs | 0.0922 | 0.273 | 0.35 | 0.723 | 0.154 [0.143 - 0.166] | 0.158 [0.138 - 0.18] |
| ALA | PUFA | Diet | Rel Abs | 0.102 | 0.435 | 0.703 | 0.574 | 0.146 [0.131 - 0.162] | 0.153 [0.128 - 0.184] |
| AA | PUFA | D5D | Rel Abs | 0.0125 | 0.328 | 0.439 | 0.723 | 0.138 [0.126 - 0.151] | 0.155 [0.139 - 0.172] |
| EPA | PUFA | D6D | Rel Abs | 0.324 | 0.0928 | 0.611 | 0.0668 | 0.12 [0.105 - 0.138] | 0.13 [0.108 - 0.156] |
| DHA | PUFA | D6D | Rel Abs | 0.419 | 0.0253 | 0.913 | 0.0334 | 0.147 [0.133 - 0.162] | 0.149 [0.128 - 0.174] |
| OEA | Acyl-EA | PLD | nM | 0.0004 | 0.647 | 0.0846 | 0.0001 | 7.31 [6.63 - 8.06] | 5.23 [4.79 - 5.7] |
| LEA | Acyl-EA | PLD | nM | 0.278 | 0.527 | 0.924 | 0.732 | 4.26 [3.9 - 4.65] | 4.5 [4.13 - 4.91] |
| aLEA | Acyl-EA | PLD | nM | 0.72 | 0.321 | 0.437 | 0.894 | 0.179 [0.161 - 0.199] | 0.184 [0.164 - 0.207] |
| DGLEA | Acyl-EA | PLD | nM | 0.002 | 0.247 | 0.919 | 0.0001 | 0.245 [0.216 - 0.276] | 0.175 [0.151 - 0.201] |
| AEA | Acyl-EA | PLD | nM | 0.0002 | 0.968 | 0.502 | 0.0001 | 2.2 [1.97 - 2.46] | 1.49 [1.36 - 1.64] |
| DEA | Acyl-EA | PLD | nM | 0.0001 | 0.791 | 0.169 | 0.0001 | 0.664 [0.571 - 0.771] | 0.378 [0.325 - 0.44] |
| DHEA | Acyl-EA | PLD | nM | 0.0003 | 0.124 | 0.173 | 0.0001 | 1.67 [1.5 - 1.86] | 1.18 [1.04 - 1.34] |

| | | | | | | | | | |
|---|---|---|---|---|---|---|---|---|---|
| POEA | Acyl-EA | PLD | Rel Abs | 0.445 | 0.954 | 0.265 | 0.0736 | 0.127 [0.111 - 0.145] | 0.119 [0.0985 - 0.144] |
| EPEA | Acyl-EA | PLD | Rel Abs | 0.621 | 0.597 | 1 | 0.0257 | 0.102 [0.0878 - 0.119] | 0.0873 [0.0704 - 0.108] |
| NO-Gly | Acyl-Gly | FAAH | nM | 0.215 | 0.401 | 0.764 | 0.0001 | 5.21 [4.8 - 5.66] | 4.7 [4.29 - 5.16] |
| NA-Gly | Acyl-Gly | FAAH | nM | 0.126 | 0.791 | 0.905 | 0.951 | 0.609 [0.533 - 0.695] | 0.72 [0.621 - 0.835] |
| 1-AG and 2-AG | MAG | PL | nM | 0.709 | 0.349 | 0.237 | 0.0089 | 25.3 [22.8 - 28] | 23.8 [21.2 - 26.8] |
| 1-LG and 2-LG | MAG | PL | nM | 0.209 | 0.456 | 0.104 | 0.849 | 260 [229 - 295] | 222 [197 - 250] |
| 1-OG and 2-OG | MAG | PL | nM | 0.0184 | 0.75 | 0.0756 | 0.789 | 409 [358 - 467] | 318 [284 - 355] |
| Ibuprofen | NSAID | Treatment | nM | 0.725 | 0.975 | 0.737 | 0.09 | 64.2 [38.6 - 107] | 41 [21.4 - 78.4] |
| Naproxen | NSAID | Treatment | nM | 0.0959 | 0.66 | 0.225 | 0.0737 | 10.3 [5.31 - 19.9] | 31.4 [7.51 - 132] |
| Acetaminophen | NSAID | Treatment | nM | 0.905 | 0.868 | 0.475 | 0.778 | 5.14 [3.09 - 8.54] | 5.06 [2.61 - 9.82] |
| Oxylipin ratios | | | | | | | | | |
| 12,13-DiHOME/EpOME | | | | 0.0017 | 0.595 | 0.302 | 0.0001 | 2.07 [1.93 - 2.22] | 1.6 [1.43 - 1.79] |
| 9_10-DiHODE/9(10)-EpODE | | | | 0.475 | 0.228 | 0.777 | 0.0002 | 1.09 [0.87 - 1.36] | 1.26 [0.861 - 1.84] |
| 13-KODE/13-HODE | | | | 0.984 | 0.333 | 0.182 | 0.0022 | 0.103 [0.0867 - 0.123] | 0.104 [0.0854 - 0.126] |
| 11,12/14,15-DiHETrE | | | | 0.0494 | 0.552 | 0.246 | 0.0029 | 0.851 [0.825 - 0.878] | 0.809 [0.771 - 0.849] |
| 14,15/11,12-DiHETrE | | | | 0.0494 | 0.552 | 0.246 | 0.0029 | 1.18 [1.14 - 1.21] | 1.24 [1.18 - 1.3] |
| Sum(DiHOME)/Sum(EpOME) | | | | 0.0156 | 0.606 | 0.519 | 0.0131 | 3.02 [2.8 - 3.26] | 2.5 [2.24 - 2.78] |
| 9,10-DiHOME/EpOME | | | | 0.0141 | 0.635 | 0.247 | 0.0567 | 11.7 [10 - 13.7] | 8.97 [7.43 - 10.8] |
| 11(12)/14(15)-EpETrE | | | | 0.636 | 0.34 | 0.0738 | 0.108 | 0.781 [0.499 - 1.22] | 0.688 [0.422 - 1.12] |
| 15_16-DiHODE/ 15(16)-EpODE | | | | 0.0315 | 0.416 | 0.257 | 0.127 | 5.43 [5 - 5.89] | 4.49 [3.98 - 5.05] |
| 12-HEPE/12-HETE | | | | 0.0303 | 0.552 | 0.278 | 0.161 | 0.137 [0.121 - 0.155] | 0.11 [0.0896 - 0.135] |
| 14_15-DiHETrE/ 14(15)-EpETrE | | | | 0.902 | 0.257 | 0.525 | 0.427 | 10.1 [7.66 - 13.4] | 10.1 [7.3 - 14.1] |
| Sum(HDoHEs) | | | | 0.0032 | 0.385 | 0.212 | 0.0001 | 3.82 [3.37 - 4.34] | 2.66 [2.19 - 3.24] |
| 17_18_DiHETE+ 19_20_DiHDoPe | | | | 0.0001 | 0.0944 | 0.645 | 0.0001 | 4.45 [3.88 - 5.11] | 7.44 [6.3 - 8.78] |
| Sum(DiHETrE/EpETrE) | | | | 0.316 | 0.387 | 0.748 | 0.102 | 20.1 [16.4 - 24.6] | 17.1 [13.1 - 22.3] |
| Sum(DiHODE)/Sum(EpODE) | | | | 0.051 | 0.537 | 0.242 | 0.272 | 5.12 [4.74 - 5.53] | 4.34 [3.86 - 4.88] |
| Bile acids | | | | | | | | | |
| CA | 1º-BA | Cyp27A1; CYP8B1 | nM | 0.0321 | 0.359 | 0.417 | 0.0406 | 27.4 [20.8 - 36.2] | 19.3 [13.6 - 27.4] |
| CDCA | 1º-BA | Cyp27A1 | nM | 0.978 | 0.212 | 0.677 | 0.158 | 38.7 [29.2 - 51.5] | 46.7 [32.6 - 66.9] |
| UDCA | 2º-BA | Microbiome | nM | 0.625 | 0.56 | 0.245 | 0.972 | 68.3 [46.5 - 100] | 90.1 [61.4 - 132] |
| DCA | 2º-BA | Microbiome | nM | 0.696 | 0.0157 | 0.316 | 0.799 | 219 [184 - 261] | 177 [120 - 259] |
| LCA | 2º-BA | Microbiome | nM | 0.411 | 0.228 | 0.797 | 0.293 | 32.2 [25.4 - 40.8] | 29.3 [23 - 37.4] |
| w-MCA | 2º-BA | Cyp27A1 | nM | 0.836 | 0.185 | 0.0573 | 0.22 | 7.34 [4.85 - 11.1] | 7.63 [5.46 - 10.7] |
| b-MCA | 2º-BA | Cyp27A1 | nM | 0.248 | 0.621 | 0.43 | 0.717 | 1.42 [0.924 - 2.2] | 1.96 [1.32 - 2.91] |
| TCA | 1º-BA Conj | BAT | nM | 0.39 | 0.594 | 0.0999 | 0.0784 | 13.9 [11 - 17.5] | 11.7 [8.1 - 16.9] |
| TCDCA | 1º-BA Conj | BAT | nM | 0.726 | 0.42 | 0.149 | 0.0007 | 28.4 [22.6 - 35.8] | 30.3 [21.6 - 42.7] |
| TUDCA | 2º-BA Conj | BAT | nM | 0.0275 | 0.777 | 0.887 | 0.0001 | 1.07 [0.78 - 1.48] | 1.92 [1.53 - 2.41] |
| TDCA | 2º-BA Conj | BAT | nM | 0.23 | 0.989 | 0.118 | 0.0019 | 12.8 [9.84 - 16.8] | 13.7 [8.95 - 21] |
| TLCA | 2º-BA Conj | BAT | nM | 0.597 | 0.947 | 0.693 | 0.259 | 9.65 [7.95 - 11.7] | 7.56 [5.18 - 11.1] |
| GCA | 1º-BA Conj | BAT | nM | 0.982 | 0.74 | 0.0467 | 0.0035 | 68.9 [56 - 84.8] | 68.6 [51.6 - 91.1] |
| GCDCA | 1º-BA Conj | BAT | nM | 0.354 | 0.584 | 0.205 | 0.0001 | 316 [260 - 384] | 367 [270 - 499] |
| GUDCA | 2º-BA Conj | BAT | nM | 0.453 | 0.607 | 0.193 | 0.0004 | 41 [32.9 - 51.2] | 46.3 [32.8 - 65.4] |
| GDCA | 2º-BA Conj | BAT | nM | 0.0653 | 0.132 | 0.146 | 0.0001 | 157 [127 - 195] | 186 [125 - 277] |
| GHDCA | 2º-BA Conj | BAT | nM | 0.228 | 0.335 | 0.936 | 0.571 | 0.891 [0.688 - 1.15] | 0.72 [0.53 - 0.979] |
| GLCA | 2º-BA Conj | BAT | nM | 0.185 | 0.281 | 0.549 | 0.0003 | 15.4 [12.5 - 19] | 21.8 [14.8 - 32.1] |
| T-a-MCA | 2º-BA Conj | BAT | nM | 0.818 | 0.778 | 0.154 | 0.135 | 2.89 [2.4 - 3.49] | 2.62 [1.9 - 3.63] |
| Bile acids ratios | | | | | | | | | |
| CA/CDCA | | | | 0.0008 | 0.788 | 0.179 | 0.0001 | 0.707 [0.571 - 0.876] | 0.363 [0.271 - 0.487] |
| GDCA/DCA | | | | 0.0031 | 0.43 | 0.405 | 0.0001 | 0.719 [0.604 - 0.856] | 1.05 [0.836 - 1.33] |
| TDCA/CA | | | | 0.0056 | 0.244 | 0.82 | 0.0001 | 0.469 [0.309 - 0.711] | 0.906 [0.517 - 1.59] |
| GCA/CA | | | | 0.0104 | 0.314 | 0.838 | 0.0001 | 2.52 [1.86 - 3.39] | 4.13 [2.84 - 6.01] |
| GDCA/CA | | | | 0.0007 | 0.664 | 0.946 | 0.0001 | 5.76 [4.01 - 8.27] | 12.5 [7.71 - 20.2] |
| TDCA/TLCA | | | | 0.0114 | 0.829 | 0.0424 | 0.0003 | 1.33 [1.1 - 1.62] | 1.81 [1.49 - 2.21] |
| (GDCA+GLCA)/(TDCA+TLCA) | | | | 0.01 | 0.0131 | 0.707 | 0.0003 | 7.04 [6.18 - 8.02] | 9.61 [7.9 - 11.7] |
| TCA/CA | | | | 0.076 | 0.223 | 0.923 | 0.0004 | 0.506 [0.358 - 0.716] | 0.711 [0.434 - 1.16] |
| TDCA/DCA | | | | 0.1 | 0.0549 | 0.448 | 0.0019 | 0.0605 [0.0467 - 0.0783] | 0.0777 [0.0556 - 0.109] |
| DCA/CA | | | | 0.0459 | 0.875 | 0.944 | 0.021 | 8.01 [5.8 - 11.1] | 12 [7.89 - 18.1] |
| GUDCA/UDCA | | | | 0.7 | 0.769 | 0.952 | 0.0584 | 0.546 [0.341 - 0.876] | 0.515 [0.334 - 0.793] |
| GLCA/CDCA | | | | 0.273 | 0.975 | 0.654 | 0.0617 | 0.397 [0.278 - 0.567] | 0.487 [0.291 - 0.817] |
| GCDCA/CDCA | | | | 0.491 | 0.519 | 0.584 | 0.0688 | 8.15 [6.21 - 10.7] | 8.36 [5.68 - 12.3] |
| GCA/GDCA | | | | 0.0679 | 0.0556 | 0.493 | 0.204 | 0.431 [0.357 - 0.519] | 0.368 [0.277 - 0.49] |
| w-MCA/UDCA | | | | 0.662 | 0.549 | 0.544 | 0.23 | 0.108 [0.0659 - 0.175] | 0.0845 [0.054 - 0.132] |
| GCDCA/GLCA | | | | 0.256 | 0.429 | 0.568 | 0.445 | 20.5 [16.2 - 26] | 16.8 [11.9 - 23.7] |
| w-MCA/T-a-MCA | | | | 0.431 | 0.353 | 0.476 | 0.529 | 3.16 [1.86 - 5.38] | 2.86 [1.82 - 4.5] |

| | | | | | | | | | |
|---|---|---|---|---|---|---|---|---|---|
| GDCA/GLCA | | | | 0.988 | 0.61 | 0.224 | 0.676 | 9.92 [8.33 - 11.8] | 8.53 [6.67 - 10.9] |
| TLCA/CDCA | | | | 0.651 | 0.674 | 0.821 | 0.734 | 0.249 [0.17 - 0.364] | 0.169 [0.0992 - 0.287] |
| T-a-MCA/CDCA | | | | 0.913 | 0.35 | 0.363 | 0.929 | 0.0747 [0.0524 - 0.106] | 0.057 [0.0351 - 0.0926] |
| (GCA+TCA)/CA | | | | 0.0148 | 0.175 | 0.839 | 0.0001 | 3.12 [2.3 - 4.23] | 5.04 [3.4 - 7.47] |
| (GLCA+TLCA)/LCA | | | | 0.719 | 0.314 | 0.473 | 0.0657 | 1.08 [0.815 - 1.44] | 1.11 [0.831 - 1.49] |
| (TCA+GCA+TDCA+GDCA)/ (GUDCA+TUDCA+GLCA+ TLCA+TCDCA+GCDCA) | | | | 0.809 | 0.456 | 0.45 | 0.705 | 0.637 [0.561 - 0.724] | 0.606 [0.511 - 0.72] |
| (TCDCA+GCDCA)/CDCA | | | | 0.479 | 0.45 | 0.549 | 0.0706 | 9.06 [6.88 - 11.9] | 9.31 [6.3 - 13.7] |
| (TUDAC+GUDCA)/UDCA | | | | 0.746 | 0.786 | 0.942 | 0.0635 | 0.589 [0.369 - 0.938] | 0.54 [0.351 - 0.832] |
| DCA/(LCA+UDCA) | | | | 0.524 | 0.396 | 0.765 | 0.522 | 1.9 [1.42 - 2.55] | 1.43 [0.971 - 2.1] |
| LCA/CDCA | | | | 0.451 | 0.585 | 0.483 | 0.859 | 0.66 [0.419 - 1.04] | 0.589 [0.42 - 0.825] |
| (TDCA+GDCA)/DCA | | | | 0.0034 | 0.207 | 0.379 | 0.0001 | 0.794 [0.664 - 0.949] | 1.17 [0.919 - 1.48] |
| UDCA/CDCA | | | | 0.691 | 0.49 | 0.847 | 0.329 | 1.39 [0.815 - 2.36] | 1.94 [1.23 - 3.07] |
| GLCA/LCA | | | | 0.721 | 0.521 | 0.601 | 0.0867 | 0.711 [0.53 - 0.955] | 0.749 [0.552 - 1.02] |
| TCDCA/CDCA | | | | 0.644 | 0.122 | 0.535 | 0.179 | 0.734 [0.53 - 1.01] | 0.687 [0.433 - 1.09] |
| TLCA/LCA | | | | 0.961 | 0.0545 | 0.71 | 0.286 | 0.282 [0.2 - 0.396] | 0.252 [0.182 - 0.347] |
| TUDCA/UDCA | | | | 0.746 | 0.949 | 0.997 | 0.234 | 2.83 [1.93 - 4.17] | 2.04 [1.33 - 3.13] |
| | | | | | Steroids | | | | |
| CRTL | Steroid | beta-HSD; Cyp1 | nM | 0.597 | 0.719 | 0.676 | 0.439 | 252 [231 - 274] | 207 [158 - 272] |
| CRTN | Steroid | 11-beta-HSD | nM | 0.188 | 0.426 | 0.294 | 0.13 | 46.6 [43.6 - 49.9] | 37.4 [29.5 - 47.3] |
| CRCTN | Steroid | Cyp11B1 | nM | 0.386 | 0.686 | 0.293 | 0.669 | 4.46 [3.84 - 5.18] | 4.9 [3.82 - 6.27] |
| 11-Deoxy-CTRL | Steroid | CYP8B1 | nM | 0.952 | 0.298 | 0.671 | 0.572 | 0.775 [0.701 - 0.857] | 0.768 [0.601 - 0.98] |
| DHEAS | Steroid | | nM | 0.0057 | 0.595 | 0.705 | 0.0002 | 2000 [1410 - 2840] | 1070 [726 - 1570] |
| TEST | Steroid | 3-beta-HSD | nM | 0.945 | 0.0016 | 0.209 | 0.0239 | 1.41 [1.1 - 1.81] | 2.5 [1.63 - 3.85] |
| 17OH-PROG | Steroid | 3-beta-HSD | nM | 0.0517 | 0.338 | 0.985 | 0.0006 | 1.04 [0.872 - 1.24] | 0.951 [0.761 - 1.19] |
| Progesterone | Steroid | Sex Hormones | nM | 0.0001 | 0.661 | 0.0876 | 0.0001 | 0.617 [0.542 - 0.701] | 0.37 [0.317 - 0.432] |
| | | | | | Steroid ratios | | | | |
| Tes/Prog | | | | 0.0021 | 0.0152 | 0.767 | 0.18 | 2.29 [1.75 - 3.01] | 6.76 [4.34 - 10.5] |
