## Supplementary material for "Association of plasma and CSF cytochrome P450, soluble epoxide hydrolase and ethanolamides metabolism with Alzheimer’s disease": Table S4

**Table S4**. The mean values and t-test and two-way ANOVA interaction p-values for all detected metabolites in **CSF**. P-values < 0.05 are colored red, p-values > 0.05 < 0.1 are colored orange.

| Metabolite information | | | | p-values | | | Mean [95%CI] Uning subjects with p of fasted >60% | |
|---|---|---|---|---|---|---|---|---|
| Metabolite name | Chemical class | Enzyme | Units | **p-value** AD vs Control | **p-value** Gender x Disease interaction | **p-value** Race x Disease interaction | Control mean [95%CI] | AD mean [95%CI] |
| Oxylipins, endocannabinoids, PUFAS and NSADs | | | | | | | | |
| PGF2a | PGs | COX2 | nM | 0.0028 | 0.0964 | 0.537 | 0.0356 [0.0313 - 0.0404] | 0.0305 [0.0278 - 0.0335] |
| F2-IsoP | PGs | Auto-ox | nM | 0.105 | 0.57 | 0.0295 | 0.269 [0.256 - 0.284] | 0.295 [0.281 - 0.309] |
| 12_13-DiHOME | vic-Diol | sEH | nM | 0.0008 | 0.435 | 0.407 | 0.0275 [0.0245 - 0.0308] | 0.0356 [0.0321 - 0.0394] |
| 9_10-DiHOME | vic-Diol | sEH | nM | 0.0003 | 0.476 | 0.161 | 0.016 [0.014 - 0.0182] | 0.0218 [0.0196 - 0.0243] |
| 15_16-DiHODE | vic-Diol | sEH | nM | 0.12 | 0.8 | 0.922 | 0.0765 [0.0667 - 0.0877] | 0.0735 [0.0662 - 0.0816] |
| 14_15-DiHETrE | vic-Diol | sEH | nM | 0.501 | 0.744 | 0.674 | 0.0461 [0.0433 - 0.0491] | 0.0473 [0.0445 - 0.0503] |
| 11_12-DiHETrE | vic-Diol | sEH | nM | 0.0315 | 0.566 | 0.477 | 0.0179 [0.0166 - 0.0193] | 0.017 [0.0158 - 0.0184] |
| 17_18-DiHETE | vic-Diol | sEH | nM | 0.34 | 0.932 | 0.942 | 0.0905 [0.0784 - 0.104] | 0.0896 [0.0776 - 0.103] |
| 14_15-DiHETE | vic-Diol | sEH | nM | 0.0022 | 0.939 | 0.661 | 0.0294 [0.0254 - 0.0339] | 0.023 [0.0195 - 0.0272] |
| 19_20-DiHDoPE | vic-Diol | sEH | nM | 0.588 | 0.128 | 0.346 | 0.0155 [0.014 - 0.0171] | 0.0176 [0.0162 - 0.0192] |
| 13-HODE | R-OH | LOX | nM | 0.67 | 0.285 | 0.244 | 1.61 [1.56 - 1.66] | 1.6 [1.56 - 1.65] |
| 9-HODE | R-OH | LOX | nM | 0.517 | 0.346 | 0.119 | 0.556 [0.535 - 0.577] | 0.542 [0.525 - 0.56] |
| 13-HOTE | R-OH | LOX | nM | 0.469 | 0.0108 | 0.583 | 0.0497 [0.0468 - 0.0527] | 0.052 [0.0482 - 0.0561] |
| 9-HOTE | R-OH | LOX | nM | 0.966 | 0.298 | 0.591 | 0.0121 [0.0113 - 0.013] | 0.0123 [0.0114 - 0.0132] |
| 12(13)-EpOME | Epox | CYP | nM | 0.0001 | 0.0414 | 0.522 | 0.215 [0.196 - 0.236] | 0.335 [0.304 - 0.369] |
| 9(10)-EpOME | Epox | CYP | nM | 0.0001 | 0.0303 | 0.584 | 0.219 [0.202 - 0.237] | 0.31 [0.277 - 0.348] |
| 20-HETE | R-OH | CYP | nM | 0.0791 | 0.539 | 0.471 | 0.138 [0.126 - 0.152] | 0.126 [0.118 - 0.135] |
| LA | PUFA | Diet | Rel Abs | 0.0001 | 0.377 | 0.581 | 0.296 [0.278 - 0.315] | 0.236 [0.224 - 0.249] |
| ALA | PUFA | Diet | Rel Abs | 0.0146 | 0.104 | 0.739 | 0.293 [0.276 - 0.31] | 0.265 [0.253 - 0.279] |
| AA | PUFA | D5D | Rel Abs | 0.0001 | 0.508 | 0.948 | 0.308 [0.287 - 0.329] | 0.247 [0.232 - 0.262] |
| EPA | PUFA | D6D | Rel Abs | 0.0001 | 0.648 | 0.444 | 0.28 [0.249 - 0.314] | 0.203 [0.184 - 0.224] |
| DHA | PUFA | D6D | Rel Abs | 0.0118 | 0.322 | 0.663 | 0.284 [0.261 - 0.308] | 0.257 [0.236 - 0.279] |
| OEA | Acyl-EA | PLD | nM | 0.0001 | 0.489 | 0.0418 | 0.126 [0.116 - 0.137] | 0.0866 [0.0813 - 0.0922] |
| LEA | Acyl-EA | PLD | nM | 0.0934 | 0.0299 | 0.655 | 0.24 [0.23 - 0.251] | 0.256 [0.246 - 0.267] |
| DHEA | Acyl-EA | PLD | nM | 0.813 | 0.469 | 0.758 | 0.0136 [0.0119 - 0.0155] | 0.016 [0.0147 - 0.0173] |
| Oxylipin ratios | | | | | | | | |
| 9(10)/12(13)-EpOME | | | | 0.0001 | 0.401 | 0.98 | 1.02 [0.972 - 1.06] | 0.928 [0.9 - 0.956] |
| 11,12/14,15-DiHETrE | | | | 0.0009 | 0.56 | | 0.389 [0.377 - 0.401] | 0.36 [0.347 - 0.373] |
| 9,10/12,13-DiHOME | | | | 0.558 | 0.593 | | 0.582 [0.532 - 0.638] | 0.613 [0.576 - 0.653] |
| 14,15/17,18-DiHETE | | | | 0.171 | 0.748 | | 0.326 [0.272 - 0.391] | 0.259 [0.214 - 0.314] |
| 12,13-DiHOME/EpOME | | | | 0.0615 | 0.875 | 0.506 | 0.128 [0.112 - 0.146] | 0.106 [0.0939 - 0.12] |
| 14,15/11,12-DiHETrE | | | | 0.0008 | 0.544 | 0.46 | 2.57 [2.5 - 2.65] | 2.78 [2.68 - 2.88] |
| 9,10-DiHOME/EpOME | | | | 0.902 | 0.656 | 0.409 | 0.0732 [0.0646 - 0.0831] | 0.0703 [0.0611 - 0.0809] |
| EPA + DHA diols | | | | 0.178 | 0.82 | 0.801 | 0.147 [0.133 - 0.161] | 0.145 [0.132 - 0.159] |
| Sum(DiHOME)/Sum(EpOME) | | | | 0.248 | 0.786 | 0.474 | 0.103 [0.0917 - 0.115] | 0.0902 [0.0795 - 0.102] |
| Sum(DiHOMEs) | | | | 0.0007 | 0.356 | 0.267 | 0.0449 [0.0402 - 0.0501] | 0.0584 [0.0529 - 0.0644] |
| Sum(EpOMEs) | | | | 0.0001 | 0.0688 | 0.585 | 0.437 [0.403 - 0.474] | 0.648 [0.584 - 0.718] |
| Sum(LA Cyp+sEH) | | | | 0.0001 | 0.0231 | 0.407 | 0.492 [0.455 - 0.531] | 0.723 [0.658 - 0.796] |
| Bile acids | | | | | | | | |
| CDCA | 1º-BA | Cyp27A1 | nM | 0.756 | 0.234 | 0.152 | 1.06 [0.887 - 1.27] | 1.23 [1.06 - 1.42] |
| UDCA | 2º-BA | Microbiome | nM | 0.425 | 0.0021 | 0.678 | 0.699 [0.58 - 0.842] | 0.886 [0.719 - 1.09] |
| DCA | 2º-BA | Microbiome | nM | 0.601 | 0.0787 | 0.216 | 1.81 [1.57 - 2.1] | 1.99 [1.67 - 2.38] |
| a-MCA | 2º-BA | Cyp27A1 | nM | 0.319 | 0.751 | 0.416 | 0.0398 [0.0317 - 0.0499] | 0.033 [0.0263 - 0.0414] |
| w-MCA | 2º-BA | Cyp27A1 | nM | 0.267 | 0.36 | 0.0969 | 0.0826 [0.0652 - 0.105] | 0.106 [0.0819 - 0.136] |
| TCA | 1º-BA Conj | BAT | nM | 0.291 | 0.164 | 0.182 | 0.405 [0.336 - 0.487] | 0.503 [0.418 - 0.605] |
| TCDCA | 1º-BA Conj | BAT | nM | 0.0552 | 0.75 | 0.303 | 0.0797 [0.0675 - 0.094] | 0.0704 [0.0603 - 0.0822] |
| TUDCA | 2º-BA Conj | BAT | nM | 0.0993 | 0.46 | 0.349 | 0.00272 [0.0022 - 0.00336] | 0.00351 [0.00293 - 0.0042] |
| TDCA | 2º-BA Conj | BAT | nM | 0.878 | 0.234 | 0.0575 | 0.0463 [0.0397 - 0.054] | 0.0459 [0.0381 - 0.0553] |
| GCA | 1º-BA Conj | BAT | nM | 0.98 | 0.277 | 0.0429 | 0.907 [0.79 - 1.04] | 0.997 [0.862 - 1.15] |
| GCDCA | 1º-BA Conj | BAT | nM | 0.267 | 0.79 | 0.0913 | 0.79 [0.691 - 0.902] | 0.808 [0.723 - 0.903] |
| GUDCA | 2º-BA Conj | BAT | nM | 0.548 | 0.872 | 0.109 | 0.0879 [0.0734 - 0.105] | 0.0992 [0.0833 - 0.118] |
| GDCA | 2º-BA Conj | BAT | nM | 0.345 | 0.17 | 0.0303 | 0.367 [0.319 - 0.421] | 0.414 [0.352 - 0.488] |

| | | | | | | | | |
|---|---|---|---|---|---|---|---|---|
| GLCA | 2º-BA Conj | BAT | nM | 0.0185 | 0.299 | 0.491 | 0.0278 [0.024 - 0.0321] | 0.0368 [0.0319 - 0.0425] |
| T-a-MCA | 2º-BA Conj | BAT | nM | 0.0262 | 0.433 | 0.463 | 0.0221 [0.0186 - 0.0264] | 0.0303 [0.0256 - 0.0358] |
| Bile acids ratios | | | | | | | | |
| (GDCA+GLCA)/(TDCA) | | | | 0.0495 | 0.578 | 0.549 | 8.88 [8.07 - 9.78] | 10.5 [9.26 - 11.9] |
| (GDCA+TDCA)/(TLCA+GLCA) | | | | 0.154 | 0.0513 | 0.0332 | 15.2 [12.8 - 18] | 13 [10.8 - 15.5] |
| (GDCA+TDCA)/(TUDCA+GUDCA) | | | | 0.981 | 0.538 | 0.991 | 4.58 [3.73 - 5.62] | 4.55 [3.66 - 5.66] |
| (TCDCA+GCDCA)/CDCA | | | | 0.241 | 0.0782 | 0.841 | 0.831 [0.707 - 0.977] | 0.727 [0.632 - 0.837] |
| (TDCA+GDCA)/DCA | | | | 0.623 | 0.274 | 0.327 | 0.233 [0.205 - 0.265] | 0.24 [0.211 - 0.272] |
| (TDCA+TCDCA)/(GDCA+GCDCA) | | | | 0.161 | 0.576 | 0.975 | 0.11 [0.101 - 0.119] | 0.0968 [0.0875 - 0.107] |
| (TUDAC+GUDCA)/UDCA | | | | 0.881 | 0.0525 | 0.347 | 0.132 [0.106 - 0.165] | 0.118 [0.0925 - 0.151] |
| DCA/(LCA+UDCA) | | | | 0.587 | 0.283 | 0.632 | 2.59 [2.15 - 3.13] | 2.25 [1.81 - 2.78] |
| GCA/GCDCA | | | | 0.161 | 0.143 | 0.291 | 1.15 [1.05 - 1.26] | 1.23 [1.13 - 1.35] |
| GCA/GDCA | | | | 0.666 | 0.671 | 0.966 | 2.47 [2.12 - 2.89] | 2.41 [2.01 - 2.88] |
| GCDCA/CDCA | | | | 0.268 | 0.0832 | 0.832 | 0.744 [0.635 - 0.872] | 0.657 [0.572 - 0.755] |
| GCDCA/GDCA | | | | 0.164 | 0.21 | 0.579 | 2.15 [1.85 - 2.51] | 1.95 [1.66 - 2.29] |
| GCDCA/GLCA | | | | 0.0029 | 0.533 | 0.0727 | 28.5 [24.2 - 33.4] | 21.9 [18.8 - 25.6] |
| GDCA/DCA | | | | 0.647 | 0.212 | 0.249 | 0.202 [0.179 - 0.229] | 0.208 [0.186 - 0.233] |
| GDCA/GLCA | | | | 0.161 | 0.0588 | 0.0376 | 13.2 [11.1 - 15.7] | 11.3 [9.37 - 13.5] |
| GLCA/CDCA | | | | 0.129 | 0.0427 | 0.0693 | 0.0262 [0.021 - 0.0326] | 0.0299 [0.0248 - 0.0362] |
| GUDCA/UDCA | | | | 0.86 | 0.0606 | 0.34 | 0.126 [0.1 - 0.158] | 0.112 [0.0874 - 0.143] |
| T-a-MCA/CDCA | | | | 0.18 | 0.122 | 0.684 | 0.0209 [0.0161 - 0.027] | 0.0246 [0.0196 - 0.031] |
| TCDCA/CDCA | | | | 0.0996 | 0.104 | 0.759 | 0.0751 [0.0608 - 0.0927] | 0.0572 [0.0469 - 0.0699] |
| TCDCA/GCDCA | | | | 0.125 | 0.512 | 0.714 | 0.101 [0.0915 - 0.111] | 0.0871 [0.0779 - 0.0974] |
| TDCA/DCA | | | | 0.708 | 0.369 | 0.458 | 0.0256 [0.0212 - 0.0308] | 0.0231 [0.0188 - 0.0283] |
| TDCA/GDCA | | | | 0.204 | 0.589 | 0.822 | 0.126 [0.114 - 0.14] | 0.111 [0.0976 - 0.126] |
| TUDCA/UDCA | | | | 0.593 | 0.0018 | 0.717 | 0.00389 [0.00296 - 0.00513] | 0.00396 [0.003 - 0.00521] |
| UDCA/CDCA | | | | 0.423 | 0.019 | 0.374 | 0.658 [0.531 - 0.816] | 0.721 [0.588 - 0.885] |
| w-MCA/T-a-MCA | | | | 0.863 | 0.184 | 0.379 | 3.73 [2.74 - 5.08] | 3.49 [2.53 - 4.81] |
| w-MCA/UDCA | | | | 0.705 | 0.265 | 0.2 | 0.118 [0.092 - 0.152] | 0.119 [0.09 - 0.158] |
| Steroids | | | | | | | | |
| CRTL | Steroid | beta-HSD; Cyp1 | nM | 0.466 | 0.798 | 0.975 | 13.4 [12.6 - 14.4] | 14.4 [13.7 - 15.2] |
| CRTN | Steroid | 11-beta-HSD | nM | 0.0048 | 0.25 | 0.378 | 3.89 [3.67 - 4.13] | 4.37 [4.16 - 4.6] |
| CRCTN | Steroid | Cyp11B1 | nM | 0.503 | 0.226 | 0.995 | 0.373 [0.338 - 0.412] | 0.407 [0.37 - 0.449] |
| 11-Deoxy-CTRL | Steroid | CYP8B1 | nM | 0.0803 | 0.0648 | 0.132 | 0.0451 [0.0414 - 0.0491] | 0.0525 [0.0484 - 0.057] |
| TEST | Steroid | 3-beta-HSD | nM | 0.49 | 0.245 | 0.827 | 0.0288 [0.0234 - 0.0355] | 0.045 [0.0368 - 0.0551] |
| 17OH-PROG | Steroid | 3-beta-HSD | nM | 0.109 | 0.408 | 0.525 | 0.0433 [0.0395 - 0.0476] | 0.0464 [0.0426 - 0.0505] |
