## Supplementary material for "Association of plasma and CSF cytochrome P450, soluble epoxide hydrolase and ethanolamides metabolism with Alzheimer’s disease": Table S5

**Table S5**. Variable importance in projection (VIP) scores for all plasma and CSF variables used for partial least square discriminant analysis (PLS-DA)

| Variable | Tissue | VIP |
|---|---|---|
| OEA/LEA | Plasam | 2.9 |
| DEA/LEA | Plasam | 2.54 |
| CSF_OEA/LEA | CSF | 2.48 |
| CSF_OEA | CSF | 2.44 |
| DHEA/LEA | Plasam | 2.18 |
| 17_18-DiHETE | Plasam | 2.16 |
| Progesterone | Plasam | 1.97 |
| CSF_12(13)-EpOME | CSF | 1.95 |
| DEA/aLEA | Plasam | 1.95 |
| DEA | Plasam | 1.79 |
| DHEAS | Plasam | 1.79 |
| OEA | Plasam | 1.77 |
| AEA | Plasam | 1.75 |
| CSF_Sum(EpOMEs) | CSF | 1.74 |
| CSF_Sum(LA Cyp+sEH) | CSF | 1.74 |
| 17_18_DiHETE+19_20_Dil | Plasam | 1.74 |
| 9-HETE | Plasam | 1.74 |
| 12,13-DiHOME/EpOME 2 | Plasam | 1.71 |
| 4-HDoHE | Plasam | 1.71 |
| DHEA/aLEA | Plasam | 1.69 |
| Tes/Prog | Plasam | 1.67 |
| CSF_9(10)/12(13)-EpOME | CSF | 1.64 |
| DHEA | Plasam | 1.64 |
| CA/CDCA | Plasam | 1.6 |
| CSF_9(10)-EpOME | CSF | 1.58 |
| 14_15-DiHETrE | Plasam | 1.55 |
| 9-HEPE | Plasam | 1.47 |
| 5-HETE | Plasam | 1.45 |
| 5-HEPE | Plasam | 1.45 |
| CSF_AA_RelAb | CSF | 1.43 |
| 12-HEPE | Plasam | 1.39 |
| Sum(HDoHEs) | Plasam | 1.38 |
| 1-OG | Plasam | 1.37 |
| CSF_GLCA | CSF | 1.36 |
| 9_12_13-TriHOME | Plasam | 1.36 |
| DGLEA | Plasam | 1.35 |
| Sum(DiHOME)/Sum(EpOI | Plasam | 1.34 |
| 11_12-DiHETrE | Plasam | 1.34 |
| CSF_PGF2a | CSF | 1.31 |
| CSF_12,13-DiHOME/EpOI | CSF | 1.3 |
| 8-HETE | Plasam | 1.3 |

| | | |
|---|---|---|
| 2-OG | Plasam | 1.29 |
| 15_16-DiHODE/15(16)-Ep( | Plasam | 1.28 |
| (GDCA+GLCA)/(TDCA+TLC/ | Plasam | 1.28 |
| 11-HETE | Plasam | 1.27 |
| CSF_GCDCA/GLCA | CSF | 1.26 |
| GDCA/CA | Plasam | 1.26 |
| CSF_LA_RelAb | CSF | 1.25 |
| 14-HDoHE | Plasam | 1.25 |
| 9_10-e-DiHO | Plasam | 1.23 |
| 12-HETE | Plasam | 1.22 |
| CSF_F2-IsoP | CSF | 1.18 |
| F2-IsoP | Plasam | 1.16 |
| 15-HEPE | Plasam | 1.16 |
| 1-LG | Plasam | 1.16 |
| CSF_(GDCA+GLCA)/(TDCA) | CSF | 1.14 |
| Sum(DiHODE)/Sum(EpODE | Plasam | 1.13 |
| CSF_UDCA | CSF | 1.12 |
| CA | Plasam | 1.12 |
| Testosterone | Plasam | 1.12 |
| GDCA/DCA | Plasam | 1.11 |
| w-MCA/UDCA | Plasam | 1.1 |
| CSF_Sum(DiHOME)/Sum(E | CSF | 1.09 |
| CSF_TEST | CSF | 1.09 |
| 15-HETE | Plasam | 1.09 |
| 2-LG | Plasam | 1.09 |
| CSF_14,15/17,18-DiHETE | CSF | 1.07 |
| (TDCA+GDCA)/DCA | Plasam | 1.07 |
| LEA | Plasam | 1.07 |
| 13-HODE | Plasam | 1.06 |
| TUDCA | Plasam | 1.06 |
| CSF_UDCA/CDCA | CSF | 1.03 |
| CSF_EPA_RelAb | CSF | 1.03 |
| PGF2a | Plasam | 1.03 |
| (w)+a+b-MCA | Plasam | 1.03 |
| AA_screen | Plasam | 1.02 |
| CSF_TCDCA/GCDCA | CSF | 1.01 |
| CSF_11-Deoxy-CTRL | CSF | 1.01 |
| LCA/CDCA | Plasam | 1.01 |
| PGE2 | Plasam | 1 |
| 12_13-DiHOME | Plasam | 0.999 |
| CSF_9_10-DiHOME | CSF | 0.986 |
| CSF_11,12/14,15-DiHETrE | CSF | 0.975 |
| CSF_14,15/11,12-DiHETrE | CSF | 0.971 |
| CSF_GUDCA/UDCA | CSF | 0.971 |
| 9,10-DiHOME/EpOME 2 | Plasam | 0.967 |
| GCA/CA | Plasam | 0.966 |
| 12-HEPE/12-HETE | Plasam | 0.964 |

| | | |
|---|---|---|
| CSF_(TUDAC+GUDCA)/UD( | CSF | 0.962 |
| 9-HODE | Plasam | 0.951 |
| (GCA+TCA)/CA | Plasam | 0.95 |
| 9(10)-EpOME | Plasam | 0.948 |
| TDCA/CA | Plasam | 0.943 |
| LA_screen | Plasam | 0.941 |
| CSF_TDCA/GDCA | CSF | 0.938 |
| GHDCA | Plasam | 0.917 |
| EPEA_Screen | Plasam | 0.912 |
| CSF_(TDCA+TCDCA)/(GDC/ | CSF | 0.881 |
| CSF_9,10-DiHOME/EpOME | CSF | 0.867 |
| GLCA | Plasam | 0.866 |
| TXB2 | Plasam | 0.86 |
| TDCA/TLCA | Plasam | 0.839 |
| (TUDAC+GUDCA)/UDCA | Plasam | 0.836 |
| PGD2 | Plasam | 0.835 |
| CSF_TCDCA | CSF | 0.799 |
| 5_6-DiHETrE | Plasam | 0.791 |
| 12_13-DiHODE | Plasam | 0.788 |
| CSF_GLCA/CDCA | CSF | 0.785 |
| CSF_GCDCA/GDCA | CSF | 0.784 |
| CSF_T-a-MCA | CSF | 0.777 |
| CSF_GDCA | CSF | 0.774 |
| CSF_DCA | CSF | 0.77 |
| CSF_20-HETE | CSF | 0.768 |
| NA-Gly | Plasam | 0.768 |
| GCA/GCDCA | Plasam | 0.766 |
| GDCA | Plasam | 0.766 |
| CSF_DHA_RelAb | CSF | 0.759 |
| CSF_GCA/GDCA | CSF | 0.758 |
| w-MCA/T-a-MCA | Plasam | 0.757 |
| CSF_9,10/12,13-DiHOME | CSF | 0.752 |
| DCA/(LCA+UDCA) | Plasam | 0.747 |
| GLCA/CDCA | Plasam | 0.733 |
| TLCA/CDCA | Plasam | 0.724 |
| ALA_screen | Plasam | 0.723 |
| 14,15/11,12-DiHETrE | Plasam | 0.72 |
| 11,12/14,15-DiHETrE | Plasam | 0.72 |
| TCA/CA | Plasam | 0.72 |
| CSF_17_18-DiHETE | CSF | 0.719 |
| CSF_13-HOTE | CSF | 0.715 |
| 2-AG | Plasam | 0.713 |
| 9_10-DiHODE/9(10)-EpOD | Plasam | 0.711 |
| GCA/GDCA | Plasam | 0.705 |
| 8_9-DiHETrE | Plasam | 0.705 |
| CSF_TCDCA/CDCA | CSF | 0.704 |
| DHA_screen | Plasam | 0.703 |

| | | |
|---|---|---|
| EPA_screen | Plasam | 0.701 |
| 15(16)-EpODE | Plasam | 0.695 |
| 5_15-DiHETE | Plasam | 0.681 |
| 11(12)-EpETrE | Plasam | 0.679 |
| UDCA/CDCA | Plasam | 0.675 |
| DCA/CA | Plasam | 0.673 |
| CSF_CRTL | CSF | 0.659 |
| NO-Gly | Plasam | 0.656 |
| 1-AG | Plasam | 0.653 |
| CSF_DHEA | CSF | 0.644 |
| CSF_DCA/(LCA+UDCA) | CSF | 0.643 |
| 17-OH Prog | Plasam | 0.634 |
| 13-KODE | Plasam | 0.621 |
| POEA_Screen | Plasam | 0.62 |
| 12(13)-EpOME | Plasam | 0.619 |
| CSF_TUDCA/UDCA | CSF | 0.616 |
| CSF_19_20-DiHDoPE | CSF | 0.616 |
| CSF_(GDCA+TDCA)/(TLCA+ | CSF | 0.613 |
| 9-HOTE | Plasam | 0.61 |
| GCDCA/CDCA | Plasam | 0.604 |
| 9_10-DiHOME | Plasam | 0.603 |
| CSF_ALA_RelAb | CSF | 0.599 |
| CSF_14_15-DiHETE | CSF | 0.588 |
| CSF_TDCA/DCA | CSF | 0.586 |
| CSF_TDCA/DCA 2 | CSF | 0.586 |
| CSF_LEA | CSF | 0.583 |
| CSF_EPA + DHA diols | CSF | 0.582 |
| CSF_CRCTN | CSF | 0.582 |
| CSF_GDCA/GLCA | CSF | 0.581 |
| CSF_CRTN | CSF | 0.58 |
| CSF_Sum(DiHOMEs) | CSF | 0.575 |
| (TCDCA+GCDCA)/CDCA | Plasam | 0.574 |
| GCDCA | Plasam | 0.574 |
| GCDCA/GLCA | Plasam | 0.573 |
| CSF_w-MCA/UDCA | CSF | 0.57 |
| 11(12)/14(15)-EpETrE | Plasam | 0.565 |
| LCA | Plasam | 0.563 |
| CSF_a-MCA | CSF | 0.562 |
| (TDCA+TCDCA)/(GDCA+GC | Plasam | 0.541 |
| (GDCA+TDCA)/(TUDCA+GL | Plasam | 0.53 |
| TLCA | Plasam | 0.528 |
| 13-HOTE | Plasam | 0.51 |
| TDCA/GDCA | Plasam | 0.508 |
| CSF_12_13-DiHOME | CSF | 0.507 |
| 9(10)-EpODE | Plasam | 0.506 |
| TCDCA/GCDCA | Plasam | 0.503 |
| CSF_DHEA/LEA | CSF | 0.496 |

| | | |
|---|---|---|
| CSF_(GDCA+TDCA)/(TUDC/ | CSF | 0.495 |
| CSF_w-MCA | CSF | 0.493 |
| Cortisone | Plasam | 0.487 |
| CSF_13-HODE | CSF | 0.477 |
| CSF_T-a-MCA/CDCA | CSF | 0.455 |
| TDCA/DCA | Plasam | 0.443 |
| CSF_TUDCA | CSF | 0.442 |
| TCA | Plasam | 0.437 |
| T-a-MCA/CDCA | Plasam | 0.436 |
| CSF_9-HODE | CSF | 0.434 |
| UDCA | Plasam | 0.397 |
| aLEA | Plasam | 0.395 |
| (GLCA+TLCA)/LCA | Plasam | 0.39 |
| w+a+(b)-MCA | Plasam | 0.373 |
| TCDCA/CDCA | Plasam | 0.371 |
| GUDCA | Plasam | 0.369 |
| CDCA | Plasam | 0.368 |
| CSF_(TCDCA+GCDCA)/CDC | CSF | 0.345 |
| 19_20-DiHDoPE | Plasam | 0.343 |
| CSF_15_16-DiHODE | CSF | 0.335 |
| GDCA/GLCA | Plasam | 0.331 |
| GCA | Plasam | 0.329 |
| CSF_GCDCA/CDCA | CSF | 0.323 |
| TDCA | Plasam | 0.323 |
| CSF_11_12-DiHETrE | CSF | 0.309 |
| Cortisol | Plasam | 0.302 |
| GCDCA/GDCA | Plasam | 0.299 |
| (GDCA+TDCA)/(TLCA+GLC/ | Plasam | 0.29 |
| CSF_GCDCA | CSF | 0.273 |
| 15_16-DiHODE | Plasam | 0.269 |
| Cortexolone | Plasam | 0.263 |
| CSF_GUDCA | CSF | 0.247 |
| (TCA+GCA+TDCA+GDCA)/( | Plasam | 0.247 |
| T-w+(a)+b-MCA | Plasam | 0.247 |
| CSF_TDCA | CSF | 0.222 |
| CSF_9-HOTE | CSF | 0.218 |
| 9_10-DiHODE | Plasam | 0.214 |
| CSF_GCA | CSF | 0.213 |
| 14_15-DiHETrE/14(15)-EpI | Plasam | 0.213 |
| CSF_CDCA | CSF | 0.196 |
| 14(15)-EpETrE | Plasam | 0.174 |
| CSF_GDCA/DCA | CSF | 0.158 |
| CSF_(TDCA+GDCA)/DCA | CSF | 0.156 |
| DCA | Plasam | 0.146 |
| TCDCA | Plasam | 0.13 |
| CSF_GCA/GCDCA | CSF | 0.122 |
| corticosterone | Plasam | 0.12 |

| | | |
|---|---|---|
| 13-KODE/13-HODE | Plasam | 0.113 |
| CSF_w-MCA/T-a-MCA | CSF | 0.111 |
| CSF_14_15-DiHETrE | CSF | 0.109 |
| CSF_17OH-PROG | CSF | 0.106 |
| CSF_TCA | CSF | 0.0631 |
| Sum(DiHETrE/EpETrE) | Plasam | 0.0621 |
