## Supplementary material for "Association of plasma and CSF cytochrome P450, soluble epoxide hydrolase and ethanolamides metabolism with Alzheimer’s disease": Table S6

**Table S6.** Stepwise logistic model predicting AD status using plasma, CSF or both plasma and CSF metabolites. Stepwise analysis was performed with the maximal validation r² as the model stopping criteria, or if an additional step increased the BIC. Model stopping point for each analysis is highlighted.

| Step | Parameter | L-R ChiSquare | Sig Prob | Entry ChiSquare | Entry "Sig Prob" | RSquare | AICc | BIC | RSquare Validation |
|---|---|---|---|---|---|---|---|---|---|
| | | | | **Plasma** | | | | | |
| 1 | DEA/LEA | 32 | 0.0001 | 27.8 | 1.36E-07 | 0.219 | 118 | 124 | 0.153 |
| 2 | OEA/LEA | 16.7 | 0.0001 | 15.1 | 1.03E-04 | 0.333 | 104 | 112 | 0.425 |
| 3 | 12,13-DiHOME/EpOME | 11.5 | 0.0007 | 10.5 | 1.21E-03 | 0.412 | 94.4 | 105 | 0.469 |
| 4 | 14_15-DiHETrE | 11.5 | 0.0007 | 11.3 | 7.95E-04 | 0.491 | 85.1 | 97.9 | 0.512 |
| 5 | 9_12_13-TriHOME | 7.07 | 0.0078 | 6.46 | 0.0111 | 0.539 | 80.2 | 95.6 | 0.559 |
| 6 | **CA/CDCA** | **4.87** | **0.0274** | **4.54** | **0.0331** | **0.572** | **77.7** | **95.4** | 0.62 |
| 7 | 13-KODE | 3.03 | 0.0817 | 3.02 | 0.0824 | 0.593 | 77 | 97.1 | 0.65 |
| | | | | **CSF** | | | | | |
| 1 | CSF_OEA/LEA | 43.7 | 0.0001 | 39.1 | 3.96E-10 | 0.154 | 244 | 251 | 0.0935 |
| 2 | CSF_12(13)-EpOME | 25.8 | 0.0001 | 24 | 9.75E-07 | 0.245 | 221 | 231 | 0.13 |
| 3 | CSF_9(10)-EpOME | 9.26 | 0.0023 | 8.86 | 0.00292 | 0.277 | 213 | 227 | 0.152 |
| 4 | **CSF_GLCA** | **7.46** | **0.0063** | **7.33** | **0.00677** | **0.304** | **208** | **224** | 0.167 |
| 5 | CSF_12_13-DiHOME | 3.72 | 0.0537 | 3.6 | 0.0577 | 0.317 | 207 | 226 | 0.183 |
| | | | | **CSF and plasma** | | | | | |
| 1 | DEA/LEA | 32 | 0.0001 | 27.8 | 1.36E-07 | 0.219 | 118 | 124 | 0.153 |
| 2 | CSF_OEA/LEA | 17.4 | 0.0001 | 16.7 | 4.36E-05 | 0.337 | 103 | 111 | 0.18 |
| 3 | DHEA/LEA | 8.11 | 0.0044 | 7.5 | 6.19E-03 | 0.393 | 97.2 | 108 | 0.284 |
| 4 | 17_18-DiHETE | 6.35 | 0.0117 | 6.29 | 1.22E-02 | 0.436 | 93 | 106 | 0.384 |
| 5 | **9_12_13-TriHOME** | **6.14** | **0.0132** | **5.77** | **0.0163** | **0.478** | **89.1** | **104** | 0.42 |
| 6 | CA/CDCA | 1.54 | 0.214 | 1.53 | 0.216 | 0.489 | 89.9 | 108 | 0.473 |
